# The results of Transcriptome-wide Mendelian Randomization (TWMR) in large-scale populations can directly validate, across scales, the results of causal inference from deep learning combined with double machine learning on single-cell transcriptomes of human samples

**DOI:** 10.64898/2026.03.16.26348532

**Authors:** Wei Ye, Xinyu Jiang, Feng (Ben) Shen

## Abstract

**Objective:** Aiming at the core problems prevalent in biomedical research, including the “translational distance”, the difficulty in aligning cross-scale studies, and the lack of direct validation of single-cell systems biology models in human samples, this study aims to verify whether the results of transcriptome-wide Mendelian randomization (TWMR) based on large-scale populations are consistent with the causal inference results of deep learning combined with double machine learning (DML) using single-cell transcriptome data from human samples, to clarify whether statistical biology and systems biology can converge to the same biological truth, and provide methodological support for mechanism dissection and precision medicine research of complex diseases such as rheumatoid arthritis (RA).

**Methods:** This study integrated multi-omics data to conduct a two-stage causal inference and cross-scale validation analysis. In the first stage, based on the summary statistics of RA genome-wide association study (GWAS) from 456,348 individuals of European ancestry in the UK Biobank (UKB), and cis-expression quantitative trait locus (cis-eQTL) data from 31,684 individuals in the eQTLGen Consortium, a two-sample Mendelian randomization approach was adopted. Transcriptome-wide causal effect analysis was performed using the inverse-variance weighted (IVW) method, MR Egger regression, and weighted median method, and gene-level causal effect values were obtained after strict quality control and multiple testing correction. In the second stage, based on single-cell RNA sequencing (scRNA-seq) data from RA patients and healthy controls (RA group: 11 samples, 211,867 cells; Healthy control group: 38 samples, 456,631 cells), after preprocessing via the Seurat pipeline, batch effect correction, and cell type annotation, a hierarchical deep neural network was constructed to complete feature compression of high-dimensional expression data, and the DML framework was used to estimate the causal effects of genes on RA disease status. Finally, Pearson correlation analysis was performed to conduct cell type-specific cross-scale validation of gene-level causal effect values obtained by the two methods, and the validated model was used to quantify the causal effects of 16 RA-related pathways from the Reactome database.

**Results:** This study confirmed that the gene causal effect values obtained from large-scale population TWMR analysis were significantly correlated with those calculated by the deep learning combined with DML model based on single-cell transcriptome data. Among them, the correlation was extremely significant (p<0.001) in core naive B cells (r=0.202, p=3.2e-05, n=414) and core naive CD4 T cells (r=0.102, p=0.037, n=412). The validated DML model successfully quantified the cell type-specific causal effect values of 16 RA-related signaling pathways.

**Conclusion:** Statistical biology and systems biology can converge to the same biological truth. The cross-scale consistency between the two can significantly shorten the “translational distance” in biomedical research, and realizes the direct validation of the single-cell systems biology causal model of human samples based on large-scale population genetic data, getting rid of the excessive dependence on animal/cell experimental models in traditional research. This research paradigm not only provides a new path for mechanism dissection and therapeutic target screening of complex diseases such as RA, but also provides a feasible solution for rare disease research to break through the limitation of GWAS sample size, and lays an important theoretical and methodological foundation for constructing standardized systems biology models of human complex diseases and promoting the development of precision medicine.

## Research Background

The multicellular “ecosystem” that underpins human health and disease is characterized by extraordinary complexity. Traditionally, this immense complexity has been addressed using reductionist research approaches, which focus on a small number of preselected study components within defined spatial and temporal scales. This strategy has led to the identification of key molecular targets for intervention in numerous human diseases, such as the mutated oncogene BRAF in melanoma, or the BCR-ABL1 gene fusion in a subset of leukemias. However, the pathogenesis of many diseases cannot be explained by a single molecular event, which poses substantial challenges for clinical intervention**^[1]^**.

Research into human diseases is confronted with the challenge of “translational distance”, defined as the discrepancy between the biological characteristics of model systems (e.g., animal experiments) and those of human tissues, which renders findings from preclinical models difficult to generalize to human populations. Non-human primates, despite being the complex models most closely related to humans, still differ from humans in numerous aspects. The unvalidated assumptions between an experimental model system and its corresponding human system constitutes the “gap” between the experimental model and human physiology, which impairs the translational potential of proposed intervention strategies — and this gap is precisely defined as “translational distance”. A core question remains: can we bridge the translational distance, or is this gap inherently insurmountable? ^[1]^

The spatiotemporal dynamics of human diseases are extraordinarily complex and span an enormous range: spatial scales extend from the nanoscale molecular level to the meter-scale whole organism level, while temporal scales range from nanosecond-scale molecular rearrangement processes to the decades-long lifespan of an individual. Take cancer as an example: multiple T-cell receptors on the surface of T cells (nanoscale) can interact with peptide-human leukocyte antigen (pMHC) complexes on the surface of antigen-presenting cells within seconds. This recognition process initiates a micron-scale tyrosine phosphorylation cascade within the T cell, which proceeds over seconds to minutes and ultimately triggers alterations in gene expression within hours. Activated T cells can migrate from the draining lymph nodes through the circulatory system to the tumor site (meter-scale); meanwhile, a subset of malignant cells can escape from the tumor, enter the bloodstream, eventually form metastatic foci, and continue to proliferate over a period of days to years. Unlike cell lines, from which samples can be collected at precisely defined time points in vitro, the collection of human samples that cover specific spatiotemporal scales poses enormous challenges. Human samples are often scarce, and it is difficult to accurately contextualize them within the full course of disease progression. ^[1]^

These issues highlight the core challenges faced by systems biology in addressing the complexity of human diseases: the translational distance, the selection of research scales, and the validation of models, among others. While emerging tools (such as single-cell analysis, spatial metabolomics, and multi-omics integration) offer potential solutions, the unification of methodological and theoretical frameworks still requires further exploration.

Our study demonstrates that statistical biology from large-scale human populations and single-cell systems biology can converge on the same biological ground truth. This finding will help narrow the translational distance — the core problem that the full complexity of human diseases cannot be faithfully recapitulated by model systems (e.g., animal experiments and in vitro cell-based assays). It also addresses the long-standing challenges of cross-scale alignment and the validation of computational models. Furthermore, systems biology models that have undergone rigorous alignment and validation can overcome the inherent limitation of statistical biology in its inability to generate predictions at the individual level, thereby facilitating the advancement of precision medicine.

## Research method

### 1. Data source

The genome-wide association study (GWAS) summary statistics for binary traits used in this study were obtained from the research published in *Nature Genetics* by Jiang et al.^[2]^ Based on the UK Biobank (UKB) resource, this study included 456,348 individuals of European ancestry. The full GWAS summary statistics for all binary traits in this study have been made publicly available and can be accessed from the official fastGWA data portal (http://fastgwa.info/ukbimpbin). The strictly calibrated test statistics and accurate estimates of allelic effects provide a reliable basis for genetic instrumental variables in the present transcriptome-wide Mendelian randomization analysis.

We obtained cis-expression quantitative trait locus (cis-eQTL) summary statistics from the eQTLGen Consortium (https://www.eqtlgen.org/), which performed the largest meta-analysis of blood-derived gene expression data to date. The dataset comprised 31,684 individuals from 37 European ancestry cohorts, providing robust estimates of genetic effects on gene expression levels. For our transcriptome-wide Mendelian randomization (TWMR) analysis, we utilized the statistically significant cis-eQTL results (filtered to require replication in at least two cohorts) and full summary statistics, which included Bonferroni-adjusted p-values for each SNP-gene pair.

Single-cell RNA sequencing (scRNA-seq) data used in this study were obtained from the research by He et al. ^[3]^ on the immunological mechanisms underlying the progression from at-risk individuals to clinical rheumatoid arthritis. The raw fastq files of scRNA-seq were deposited in the dbGap database (Accession No.: phs003944.v1.p1). Processed scRNA-seq data were available from the Gene Expression Omnibus (GEO) database (Accession No.: GSE274680). The rheumatoid arthritis (RA) group contained 11 samples with a total of 211,867 cells, and the healthy control group contained 38 samples with a total of 456,631 cells.

### 2. Methods of Mendelian Randomization Analysis

We performed a two-sample Mendelian randomization (MR) analysis to investigate the causal associations between gene expression (exposure) and rheumatoid arthritis (RA, outcome). All statistical analyses were conducted using R software (version ≥4.0.0), with core packages including TwoSampleMR, dplyr, tidyr, foreach.

#### 2.1 Data Sources and Preprocessing

Exposure data (eQTL): The effect size (β) and standard error (SE) for each SNP were calculated as: 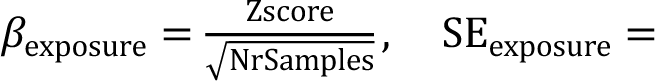 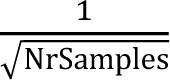 Outcome data (RA GWAS): Summary statistics of RA genome-wide association study (GWAS, accession: GCST90044540, build GRCh37) were obtained. Extracted SNPs overlapping with the exposure dataset.

#### 2.2 Data Harmonization and Quality Control

The harmonise_data() function (with action = 2) was used to align the allele directions between exposure and outcome datasets, removing SNPs with inconsistent allele information. To ensure robust instrumental variable (IV) analysis, only genes with at least 3 valid SNPs were retained for subsequent MR analysis.

#### 2.3 MR Analysis

For each eligible gene, we performed MR analysis with three complementary methods:

Inverse-Variance Weighted (IVW) (primary method, assuming no horizontal pleiotropy);

MR Egger regression (robust to horizontal pleiotropy, with intercept testing for pleiotropy);

Weighted median (valid when ≥50% of IVs are valid).

We further assessed heterogeneity across SNPs using the mr_heterogeneity() function (Q statistic and corresponding P-value) and horizontal pleiotropy using the MR Egger intercept test (mr_pleiotropy_test()).

#### 2.4 Randomly select 600 genes as the target genes for single-cell model analysis

6,000 results from the Transcriptome-wide Mendelian Randomization (TWMR) analysis (after excluding heterogeneity and pleiotropy) were used to randomly select 600 genes for the subsequent deep learning and doubly robust machine learning analysis of single-cell transcriptomic data.

### 3. Methods for Causal Inference Using Deep Learning and Double Machine Learning on Single-Cell Transcriptomic Data

#### 3.1 Study Design and Data Acquisition

Single-cell RNA sequencing (scRNA-seq) data were divided into two groups: the patient group (RA) consisting of rheumatoid arthritis patients and the control group (CON1) including healthy individuals.

##### 3.2.1 Data Loading and Initial Processing

scRNA-seq data were imported and processed using the Seurat v4 package in R. The dataset was normalized using the ‘LogNormalizè method with a scaling factor of 10,000 to account for differences in sequencing depth across cells:

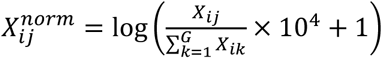

where 𝑋_𝑖𝑗_ represents the raw read count of gene 𝑗 in cell 𝑖, 𝐺 denotes the total number of genes, and 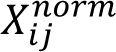 is the normalized expression value.

##### 3.2.2 Variable Feature Selection and Expression Scaling

Variable features (highly variable genes, HVGs) were identified using the variance stabilizing transformation (VST) method, with the top 4,096 HVGs retained to capture biological heterogeneity while reducing computational complexity. Expression values of HVGs were scaled to have a mean of 0 and standard deviation of 1 to eliminate technical biases:

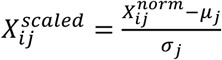

where 𝜇_𝑗_ and 𝜎_𝑗_ represent the mean and standard deviation of normalized expression values for gene 𝑗 across all cells, respectively.

#### 3.3 Batch Effect Correction and Cell Type Annotation

##### 3.3.1 Dimensionality Reduction and Batch Effect Correction

Principal Component Analysis (PCA) was performed on scaled HVG expression matrices to reduce dimensionality to 50 principal components (PCs). Batch effects across individual samples were corrected. Uniform Manifold Approximation and Projection (UMAP) was applied to the corrected PCA space for non-linear dimensionality reduction, followed by graph-based clustering with a resolution of 0.8 to identify cell clusters.

##### 3.3.2 Cell Type Annotation

Cell type annotation was conducted using a marker gene-based scoring approach. A panel of lineage-specific marker genes was defined for 16 major immune cell types ((core naive CD4+ T cells, central memory (CM) CD4+ T cells, GZMB+ CD27-effector memory (EM) CD4+ T cells, GZMB+ CD27+ EM CD4+ T cells, SOX4+ naive CD4+ T cells, ISG+ naive CD4+ T cells, T follicular helper (TFH)-like CD4+ T cells, naive CD8+ T cells, effector CD8+ T cells), natural killer (NK) cells, and an unannotated unknown). For each cell type, a signature score was calculated as the mean normalized expression of its marker genes:

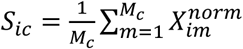

where 𝑆_𝑖𝑐_ is the signature score of cell 𝑖 for cell type 𝑐, 𝑀_𝑐_ denotes the number of valid marker genes for cell type 𝑐, and 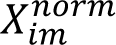 is the normalized expression of marker gene 𝑚 in cell 𝑖. Cells were assigned to the cell type with the highest signature score (threshold = 0.5); cells with scores below the threshold were labeled as “Unknown”. Only cell types with ≥100 cells were retained for subsequent analysis.

#### 3.4 Deep Learning-Based Feature Compression

##### 3.4.1 Global Feature Compression for Background Genes

A hierarchical deep neural network was constructed to perform unsupervised feature compression on background genes (HVGs excluding target genes and pathway genes). The network architecture consisted of an input layer, an 8-layer encoder module for dimensionality reduction, and a symmetric decoder module for reconstruction:

- Input layer: dimensionality matching the number of background genes (up to 4,096)

- Encoder module (sequential dimensional reduction): 4096 → 2048 → 1024 → 512 → 256 → 128 → 64 → 32 (bottleneck layer)

- Each encoder layer was followed by ReLU activation, 30% dropout for regularization, and batch normalization to stabilize training

For 12 epochs using mean squared error (MSE) as the loss function and Adam optimizer (learning rate = 0.0005). Early stopping (patience = 2) and learning rate reduction on plateau (factor = 0.5, patience = 1) were applied to prevent overfitting. The bottleneck layer output (32-dimensional latent representation) was extracted as the compressed global feature matrix (𝑋) for subsequent analysis.

##### 3.4.2 Pathway-Specific Feature Compression

For pathway-related genes, a dedicated deep neural network was constructed to compress the high-dimensional pathway gene expression matrix into a 1-dimensional latent representation.

#### 3.5 Double Machine Learning (DML) Analysis

DML Framework for Causal Effect Estimation

Double Machine Learning was applied to estimate the causal effect of target genes/pathway features on disease status (RA vs. Control). With the disease status (𝑌) coded as 1 (RA) or 0 (Control). For each target gene (expression 𝑇) or pathway feature (1-dimensional latent representation 𝑇), the DML framework consisted of two stages:

Stage 1: Residual Calculation

- Treatement model: Regress 𝑇 on the compressed global feature matrix 𝑋 to obtain residuals𝑇_𝑟𝑒𝑠_:

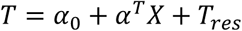

- Outcome model: Regress 𝑌 on 𝑋 using binomial generalized linear model (GLM) to obtain predicted values 𝑌^ and residuals𝑌_𝑟𝑒𝑠_:

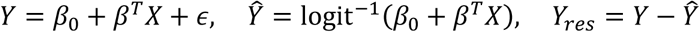

Stage 2: Causal Effect Estimation

Regress the outcome residuals 𝑌_𝑟𝑒𝑠_ on the treatment residuals 𝑇_𝑟𝑒𝑠_ to estimate the causal effect size 𝜃:

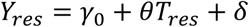

### 4. Model Validation

#### Correlation analysis was performed to validate the model for single-cell transcriptomics data constructed with deep learning and double machine learning

Stratified by each cell type, Pearson correlation analysis was conducted to compare the gene-level θ values generated from the single-cell transcriptomics data using the deep learning and double machine learning pipeline against the gene-level b values estimated by Transcriptome-wide Mendelian Randomization (TWMR). The number of matched genes included in the correlation analysis was tabulated.

### 5. Application of the Validated Model

We applied the validated single-cell transcriptomics model constructed with deep learning and double machine learning to calculate the theta (θ) values of rheumatoid arthritis (RA)-related signaling pathways.

First, a total of 16 RA-related signaling pathways were retrieved from the Reactome database (https://reactome.org/), and the biological macromolecules involved in each pathway are detailed in the Supplementary Materials. Subsequently, the θ value for each of the above pathways was computed using the aforementioned validated deep learning and double machine learning model based on single-cell transcriptomics data.

## Research results

The b-values of causal effects derived from transcriptome-wide Mendelian randomization (TWMR) analysis are detailed in Figure 1.

**Figure 1.**
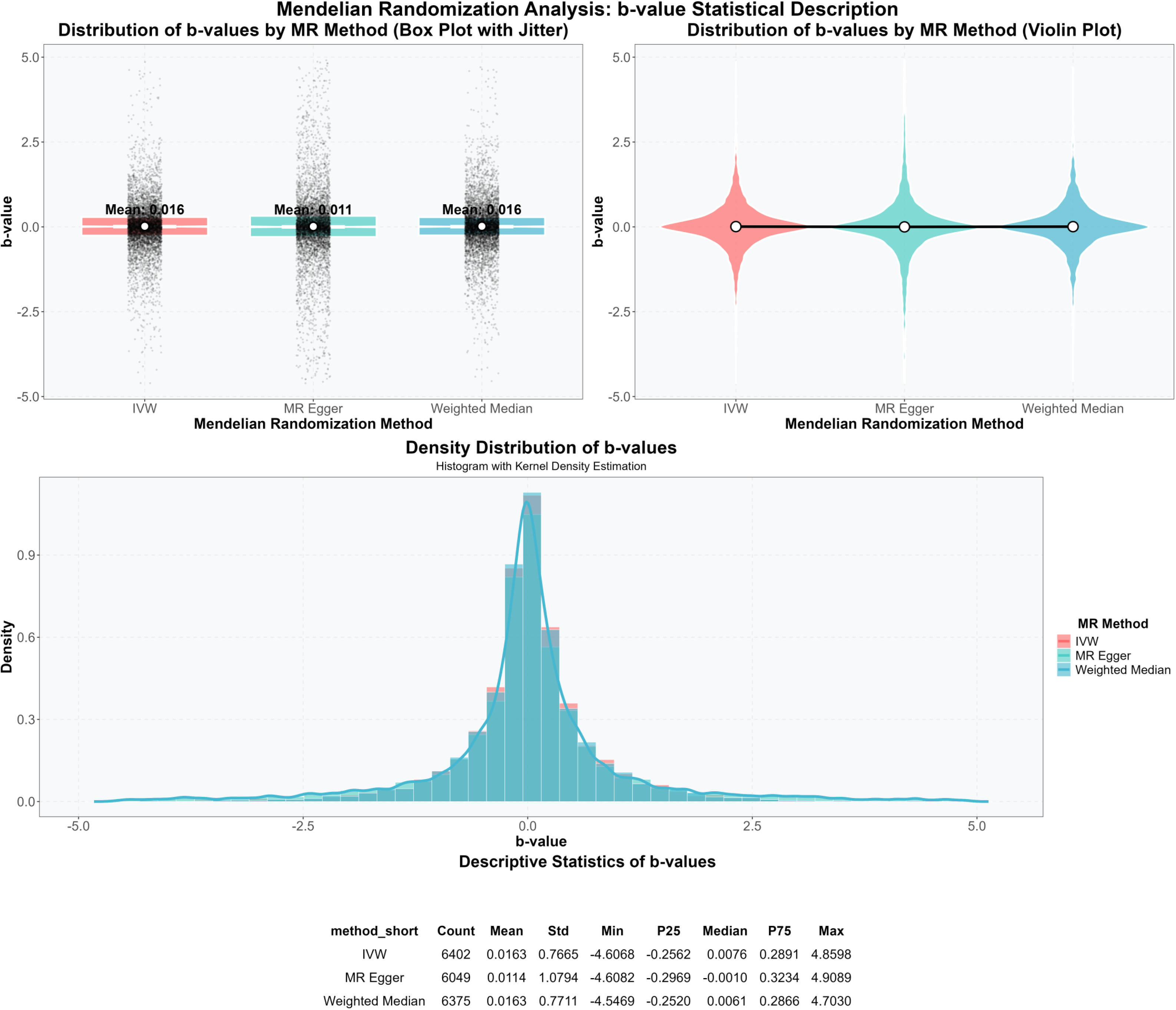
Statistical description of causal effect b-values from transcriptome-wide Mendelian randomization (TWMR) analysis in the full gene set. (Top left) Box plot with jitter showing the distribution of b-values estimated by three MR methods: inverse-variance weighted (IVW), MR Egger regression, and weighted median. (Top right) Violin plot visualizing the probability density distribution of b-values across the three MR methods. (Middle) Histogram with kernel density estimation showing the overall density distribution of b-values from the three MR methods. (Bottom) Descriptive statistics table of b-values, including gene count, mean, standard deviation (Std), minimum, 25th percentile (P25), median, 75th percentile (P75), and maximum for each MR method.

The b-values of causal effects obtained from TWMR analysis in the 600-gene subset for single-cell validation are detailed in Figure 2

**Figure 2.**
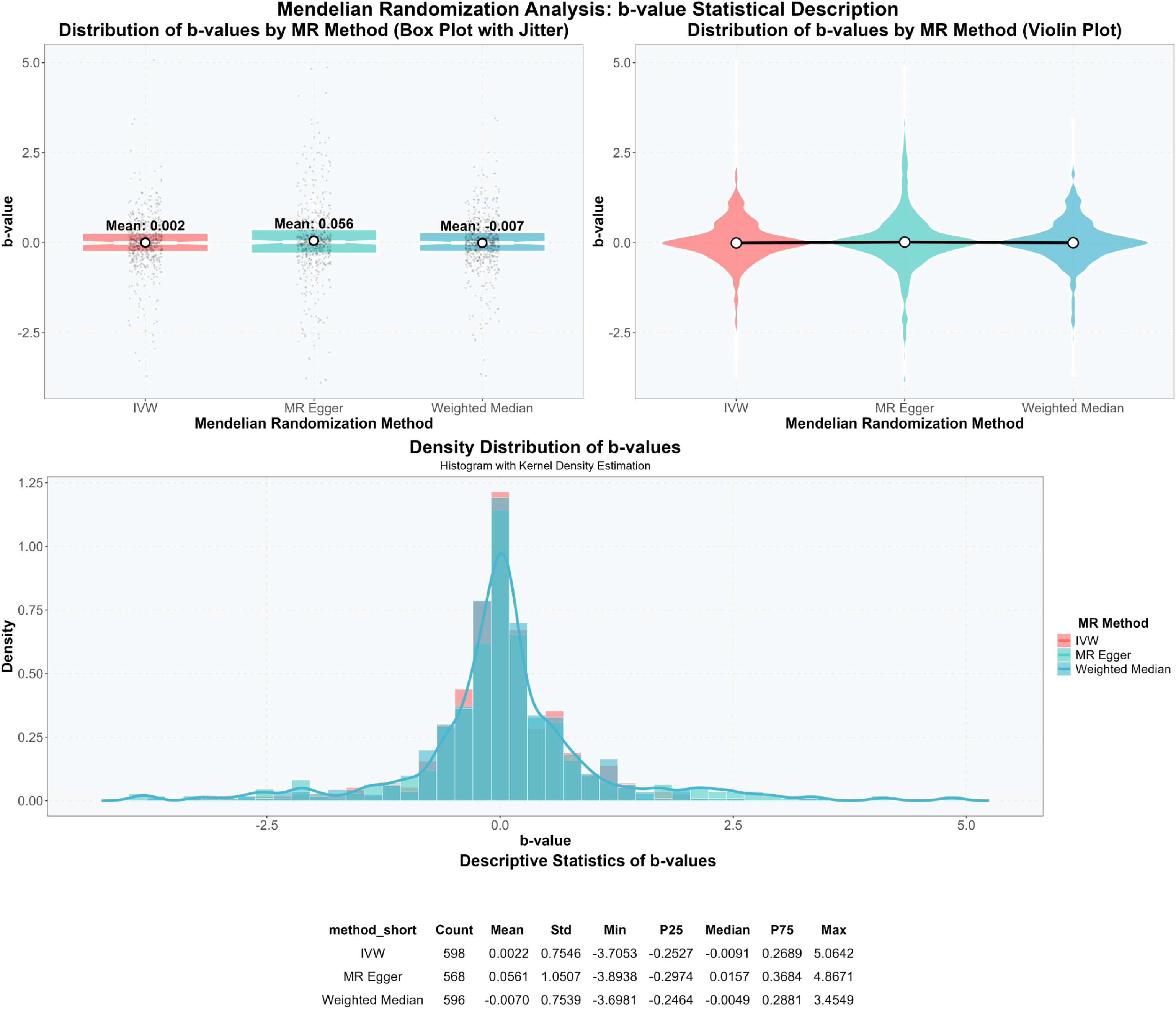
Statistical description of causal effect b-values from TWMR analysis in the 600-gene subset for single-cell validation. (Top left) Box plot with jitter showing the distribution of b-values estimated by IVW, MR Egger regression, and weighted median methods in the randomly selected 600-gene subset. (Top right) Violin plot visualizing the probability density distribution of b-values across the three MR methods in the 600-gene subset. (Middle) Histogram with kernel density estimation showing the overall density distribution of b-values from the three MR methods in the 600-gene subset. (Bottom) Descriptive statistics table of b-values for the 600-gene subset, including gene count, mean, standard deviation (Std), minimum, 25th percentile (P25), median, 75th percentile (P75), and maximum for each MR method.

**Figure 3.**
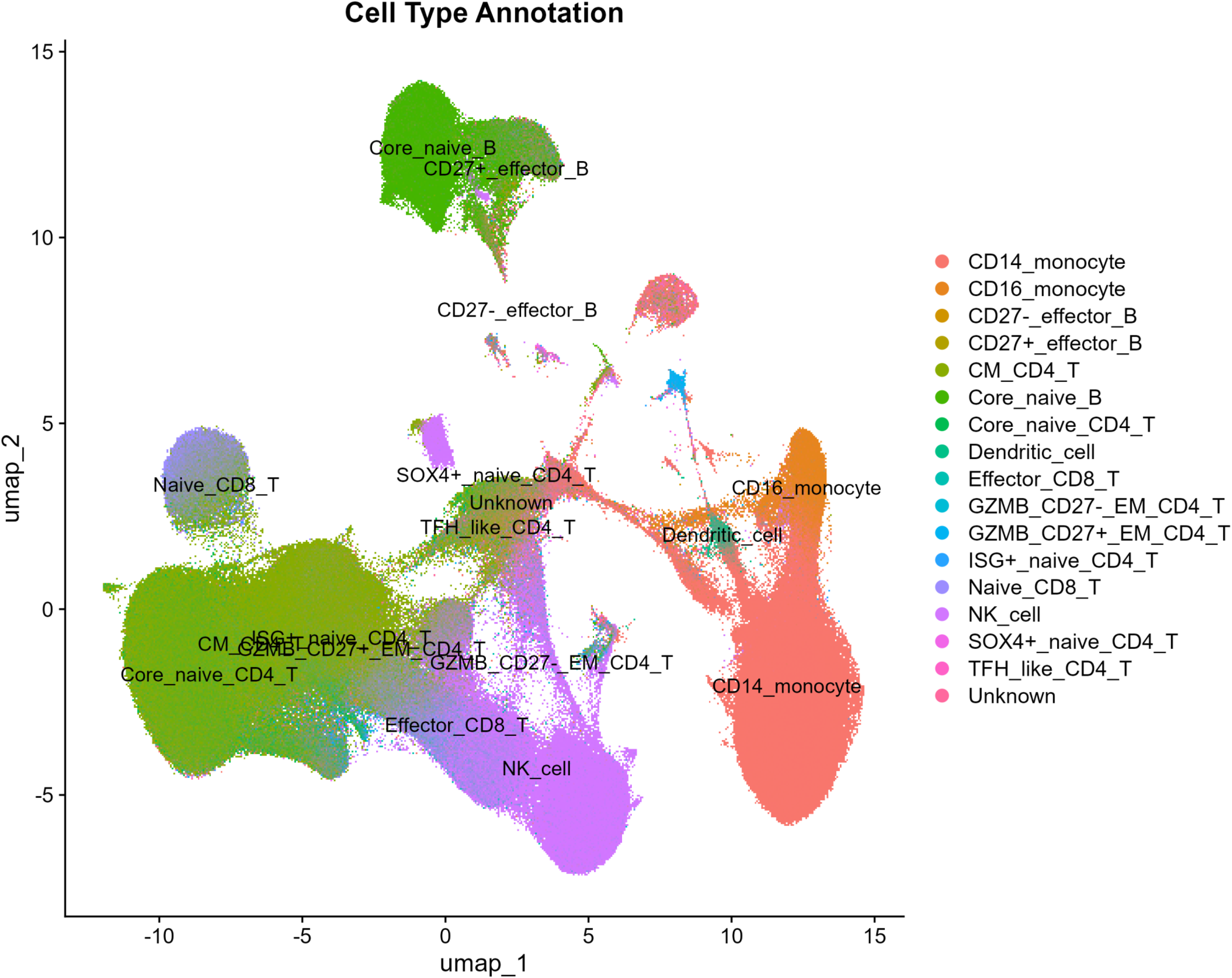
UMAP visualization of single-cell transcriptome clustering and cell type annotation. This Uniform Manifold Approximation and Projection (UMAP) plot displays the low-dimensional embedding of the single-cell RNA sequencing (scRNA-seq) dataset, where each point corresponds to an individual cell. Cells are color-coded according to their annotated cell types, with the names of major immune cell populations directly labeled on the plot. A total of 16 distinct cell populations were identified, including myeloid subsets (CD14+ monocytes, CD16+ monocytes, dendritic cells), B lymphocyte subsets (core naive B cells, CD27+ effector B cells, CD27- effector B cells), T lymphocyte subsets (core naive CD4+ T cells, central memory (CM) CD4+ T cells, GZMB+ CD27- effector memory (EM) CD4+ T cells, GZMB+ CD27+ EM CD4+ T cells, SOX4+ naive CD4+ T cells, ISG+ naive CD4+ T cells, T follicular helper (TFH)-like CD4+ T cells, naive CD8+ T cells, effector CD8+ T cells), natural killer (NK) cells, and an unannotated unknown cell population.

**Figure 4.**
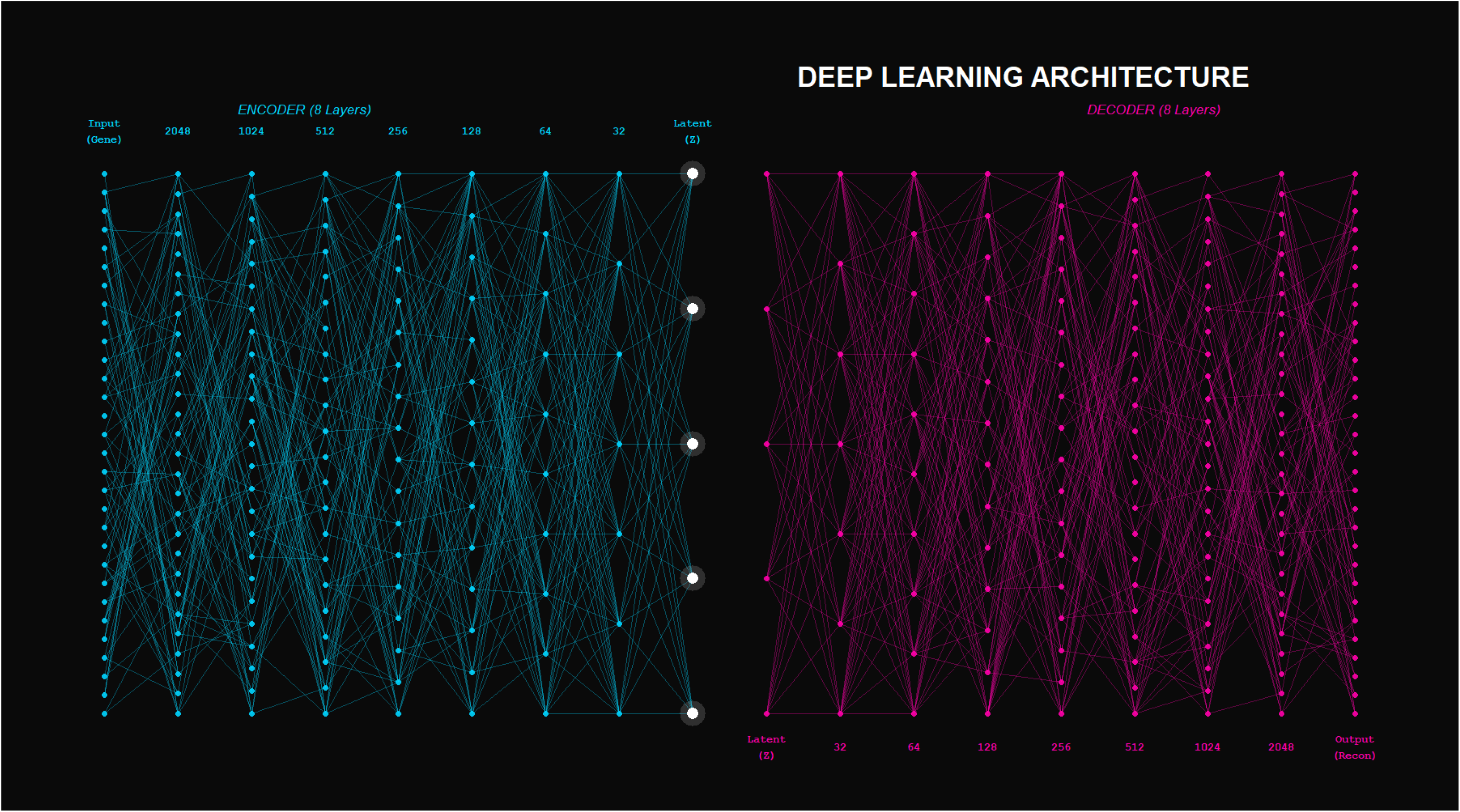
Architecture of the Hierarchical Deep Neural Network for unsupervised feature compression of single-cell transcriptomic data. The model consists of an 8-layer encoder module for sequential dimensionality reduction and a symmetric 8-layer decoder module for data reconstruction. The input layer matches the dimensionality of the background gene expression matrix (up to 4096 highly variable genes), the encoder compresses the input into a 32-dimensional latent representation (bottleneck layer, Z), and the decoder reconstructs the original gene expression input from the latent space. Each encoder layer is followed by ReLU activation, dropout regularization, and batch normalization.

**Figure 5.**
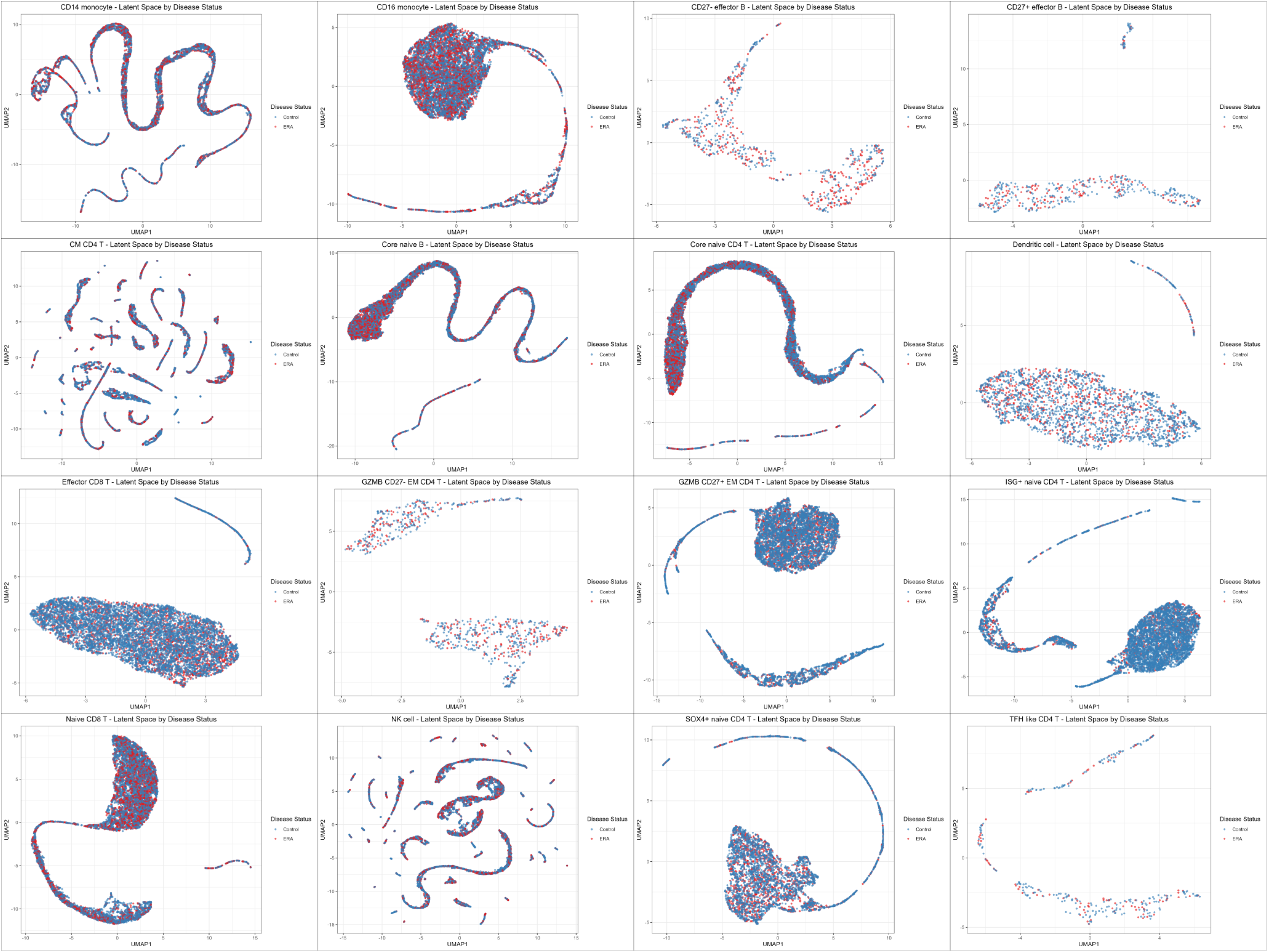
UMAP latent space embeddings of individual immune cell subsets stratified by disease status. Each panel shows the Uniform Manifold Approximation and Projection (UMAP) visualization of the latent space for a single immune cell population, with cells colored by disease status (blue: Control group, red: ERA group). Plots are generated for 16 distinct immune cell subsets, including CD14 monocytes, CD16 monocytes, CD27⁻ effector B cells, CD27⁺ effector B cells, central memory (CM) CD4 T cells, core naive B cells, core naive CD4 T cells, dendritic cells, effector CD8 T cells, GZMB⁻ effector memory (EM) CD4 T cells, GZMB⁺ effector memory (EM) CD4 T cells, ISG⁺ naive CD4 T cells, naive CD8 T cells, natural killer (NK) cells, SOX4⁺ naive CD4 T cells, and T follicular helper (TFH)-like CD4 T cells.

**Figure 6.**
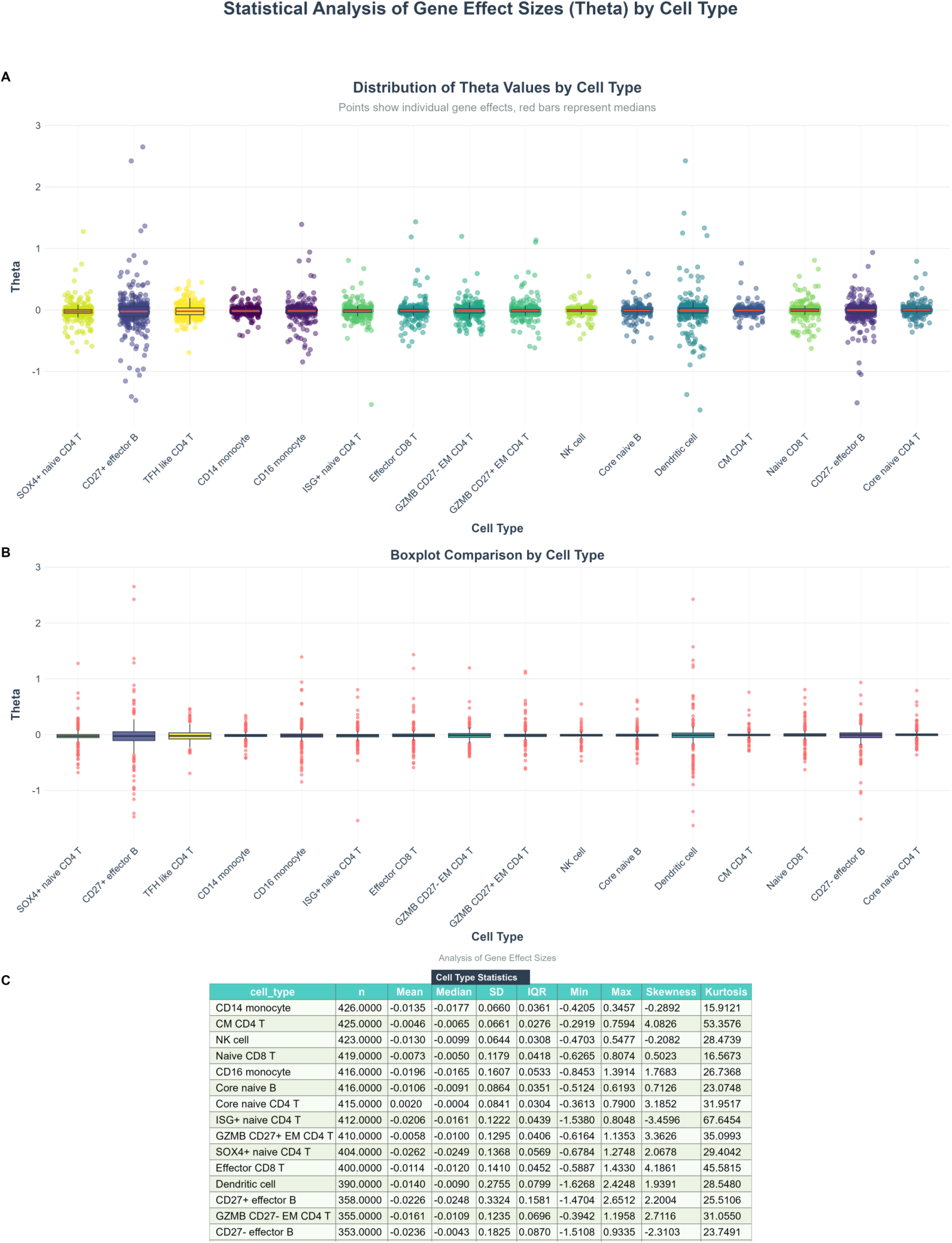
Statistical analysis of gene-level causal effect sizes (Theta values) estimated by the DML model across immune cell types. (A) Distribution of Theta values across cell types, with individual points representing the causal effect of a single gene and error bars indicating the median Theta value for each cell type. (B) Box plot comparison of Theta value distributions across all analyzed immune cell types. (C) Statistical summary table of Theta values for each cell type, including the number of matched genes (n), mean, median, standard deviation (SD), interquartile range (IQR), minimum, maximum, skewness, and kurtosis.

**Figure 7.**
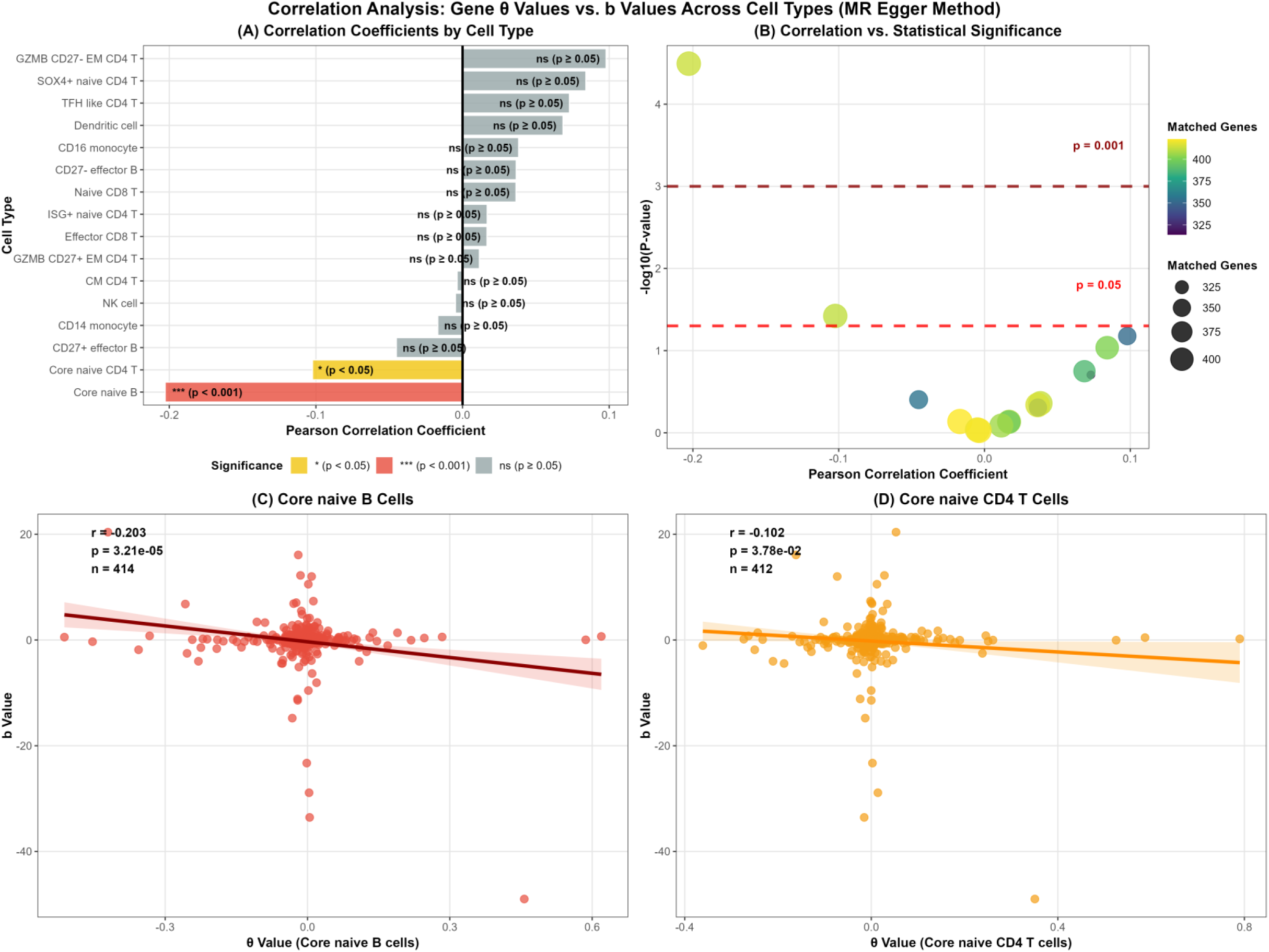
Cross-scale Pearson correlation analysis between TWMR b-values (MR Egger regression method) and single-cell DML Theta values across immune cell types. (A) Pearson correlation coefficients for gene-level causal effect estimates between TWMR (MR Egger regression method) and the single-cell DML model, stratified by cell type. (B) Correlation strength versus statistical significance across cell types, with significant correlations (p < 0.01) marked with ** , with significant correlations (p < 0.05) marked with * and non-significant results (p ≥ 0.05) labeled as ns. (C) Scatter plot of gene-level b-values (MR Egger regression method) versus Theta values in core naive B cells, with the Pearson correlation coefficient (r), p-value, and number of matched genes (n) annotated. (D) Scatter plot of gene-level b-values (MR Egger regression method) versus Theta values in core naive CD4 T cells, with the Pearson correlation coefficient (r), p-value, and number of matched genes (n) annotated.

**Figure 8.**
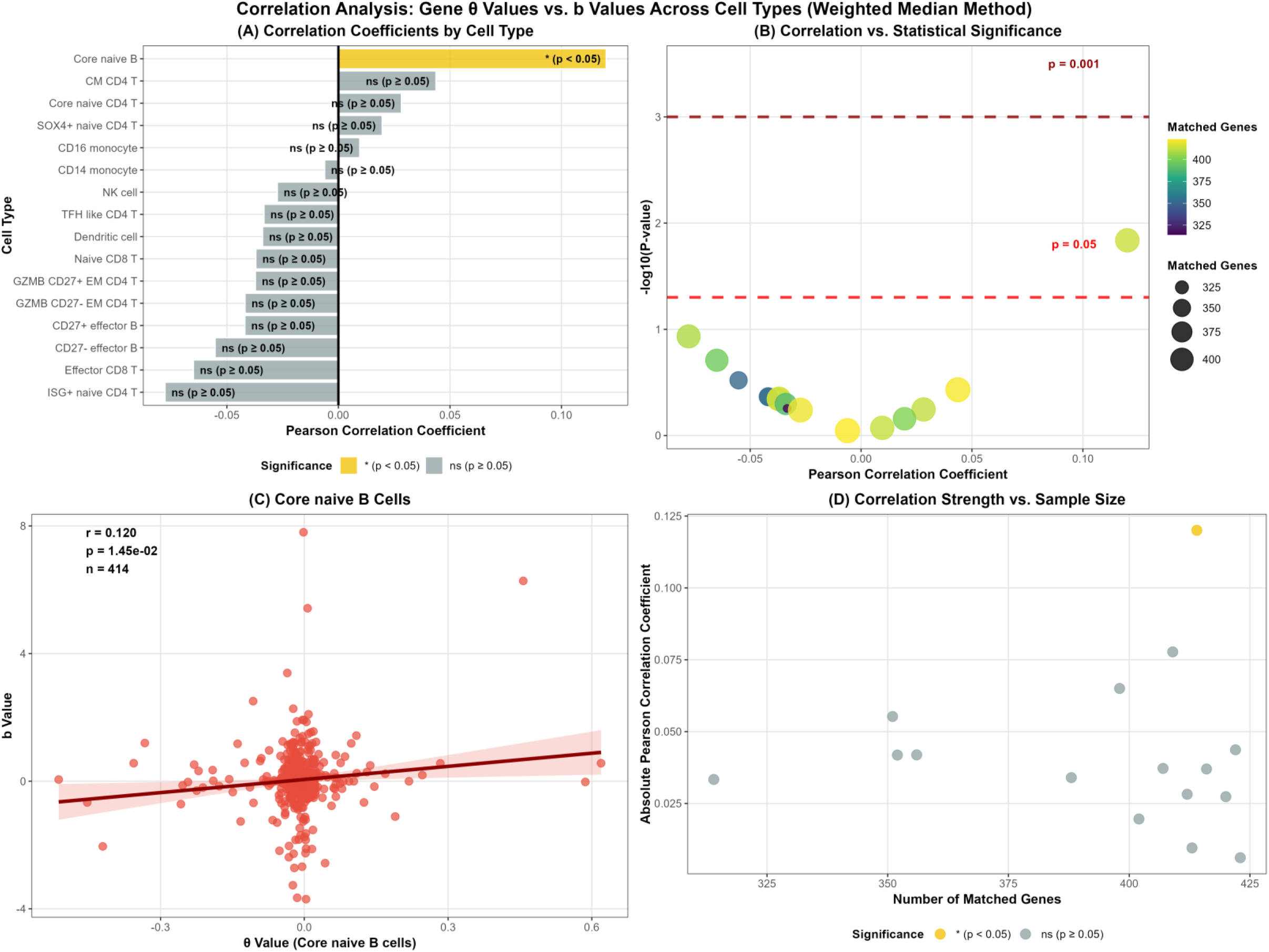
Cross-scale Pearson correlation analysis between TWMR b-values (weighted median method) and single-cell DML Theta values across immune cell types. (A) Pearson correlation coefficients for gene-level causal effect estimates between TWMR (weighted median) and the single-cell DML model, stratified by cell type. (B) Correlation strength versus statistical significance across cell types, with significant correlations (p < 0.01) marked with ** , with significant correlations (p < 0.05) marked with * and non-significant results (p ≥ 0.05) labeled as ns. (C) Scatter plot of gene-level b-values (weighted median) versus Theta values in core naive B cells, with the Pearson correlation coefficient (r), p-value, and number of matched genes (n) annotated. (D) Scatter plot (D) showing the association between absolute Pearson correlation coefficient (y-axis, correlation strength) and number of matched genes (x-axis, sample size).

**Figure 9.**
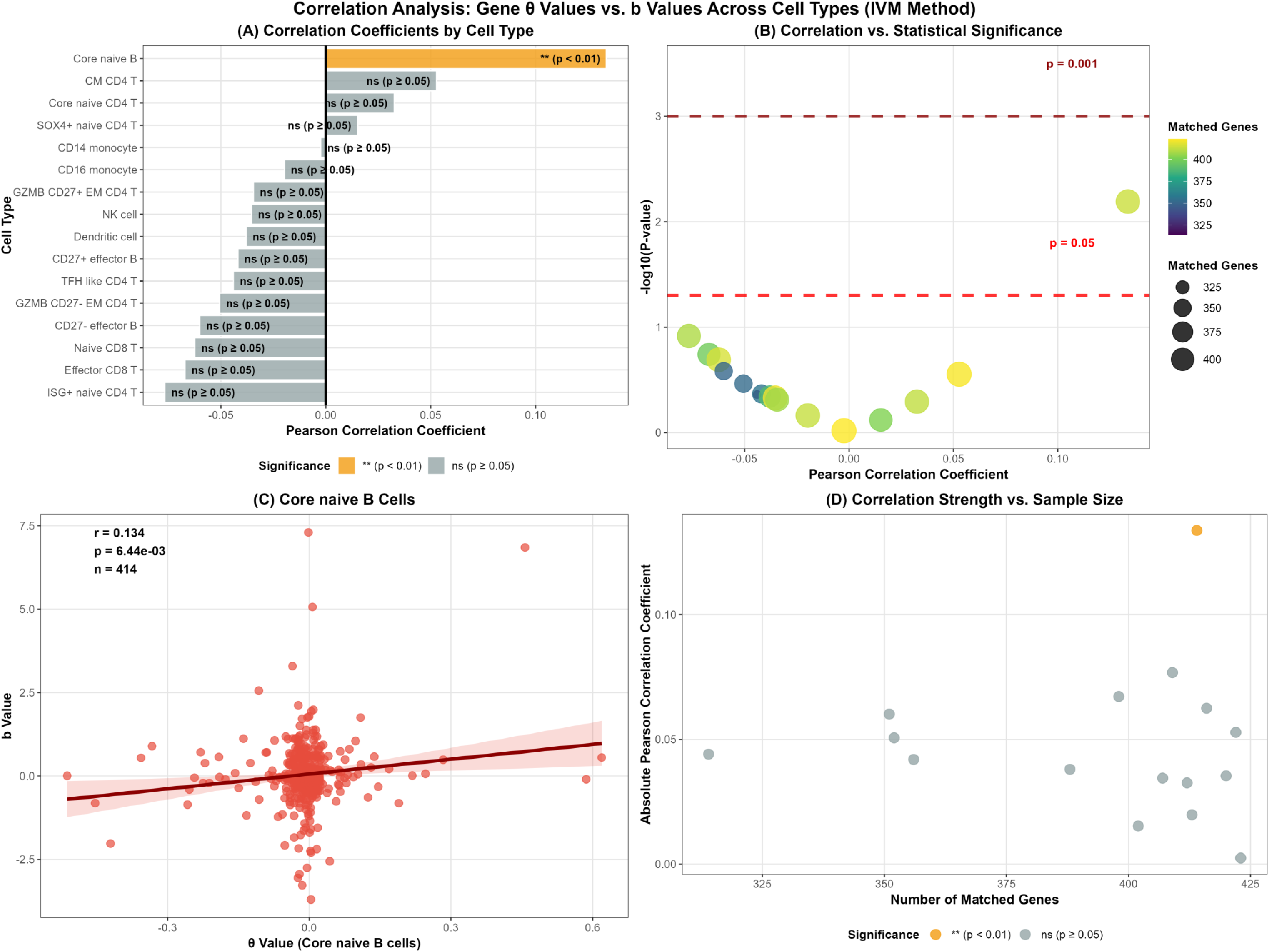
Cross-scale Pearson correlation analysis between TWMR b-values (inverse-variance weighted method) and single-cell DML Theta values across immune cell types. (A) Pearson correlation coefficients for gene-level causal effect estimates between TWMR (inverse-variance weighted) and the single-cell DML model, stratified by cell type. (B) Correlation strength versus statistical significance across cell types, with significant correlations (p < 0.01) marked with ** , with significant correlations (p < 0.05) marked with * and non-significant results (p ≥ 0.05) labeled as ns. (C) Scatter plot of gene-level b-values (inverse-variance weighted) versus Theta values in core naive B cells, with the Pearson correlation coefficient (r), p-value, and number of matched genes (n) annotated. (D) Scatter plot (D) showing the association between absolute Pearson correlation coefficient (y-axis, correlation strength) and number of matched genes (x-axis, sample size).

**Figure 10.**
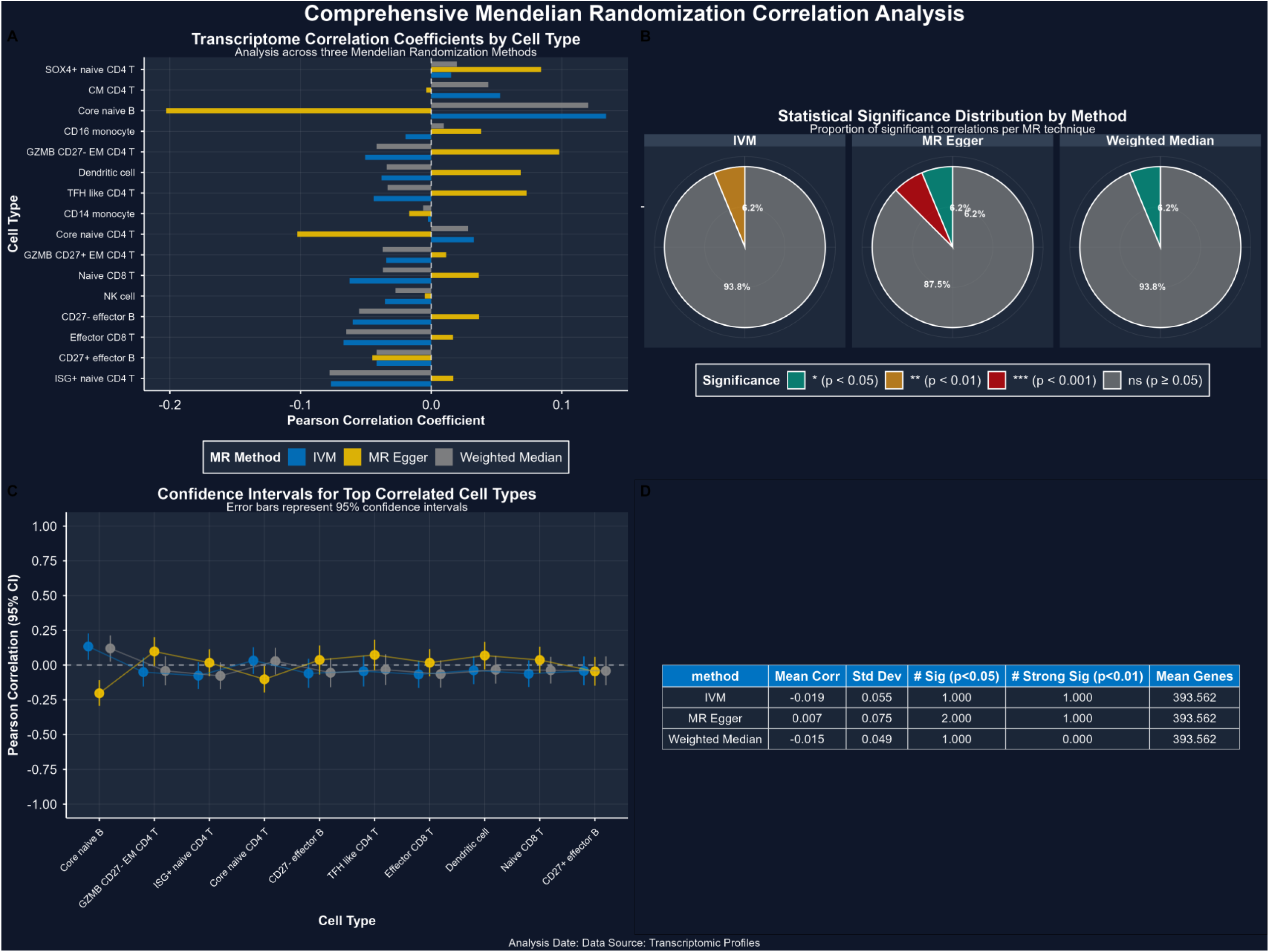
Comprehensive Mendelian randomization (MR) correlation analysis of transcriptomic profiles across immune cell types. This multi-panel figure presents transcriptome-wide association analysis evaluated via three complementary MR methods: inverse variance weighted (IVM), MR Egger, and Weighted Median. (A) Horizontal bar plot showing Pearson correlation coefficients for each immune cell type, estimated by the three MR methods. (B) Pie charts illustrating the distribution of statistical significance for MR results, stratified by analytical method. Significance thresholds are defined as: *p < 0.05 (teal), **p < 0.01 (orange), ***p < 0.001 (red), and non-significant (ns, p ≥ 0.05, gray). (C) Scatter plot with error bars depicting Pearson correlation coefficients and their corresponding 95% confidence intervals for the top correlated immune cell types across the three tested MR methods. (D) Summary statistics table for the three MR methods, including mean correlation coefficient, standard deviation (Std Dev), number of significant results at p < 0.05, number of strongly significant results at p < 0.01, and mean number of genes included in the analysis.

**Figure 11.**
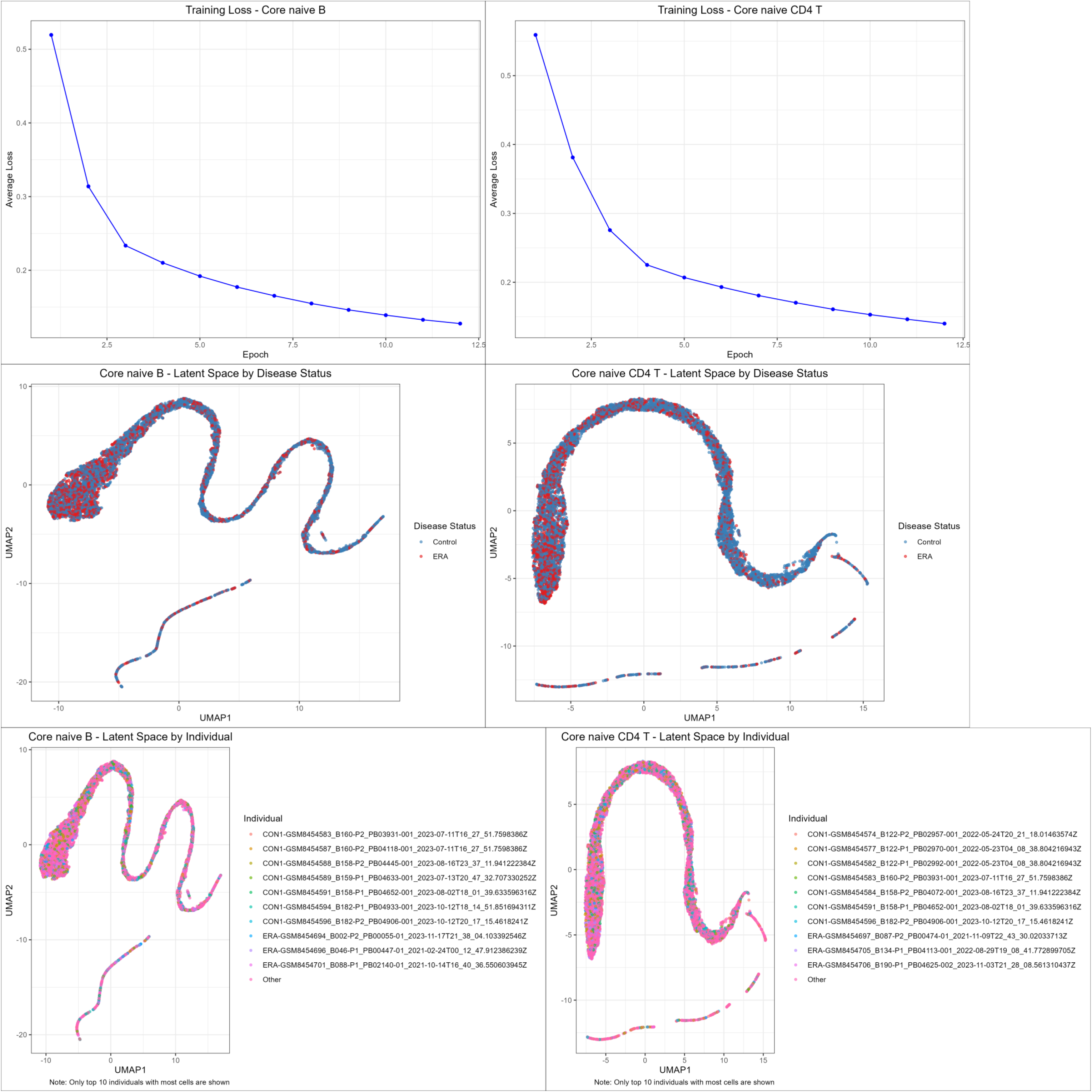
Training performance and latent space visualization of the Hierarchical Deep Neural Network model. (Top row) Training loss curves of the Hierarchical Deep Neural Network across 12 epochs for core naive B cells (left) and core naive CD4+ T cells (right), with mean squared error (MSE) as the loss function. (Middle row) UMAP visualization of the 32-dimensional bottleneck latent space, colored by disease status (RA vs. healthy control) for core naive B cells (left) and core naive CD4+ T cells (right). (Bottom row) UMAP visualization of the 32-dimensional bottleneck latent space, colored by individual sample for core naive B cells (left) and core naive CD4+ T cells (right); only the top 10 individuals with the highest cell counts are shown.

**Figure 12.**
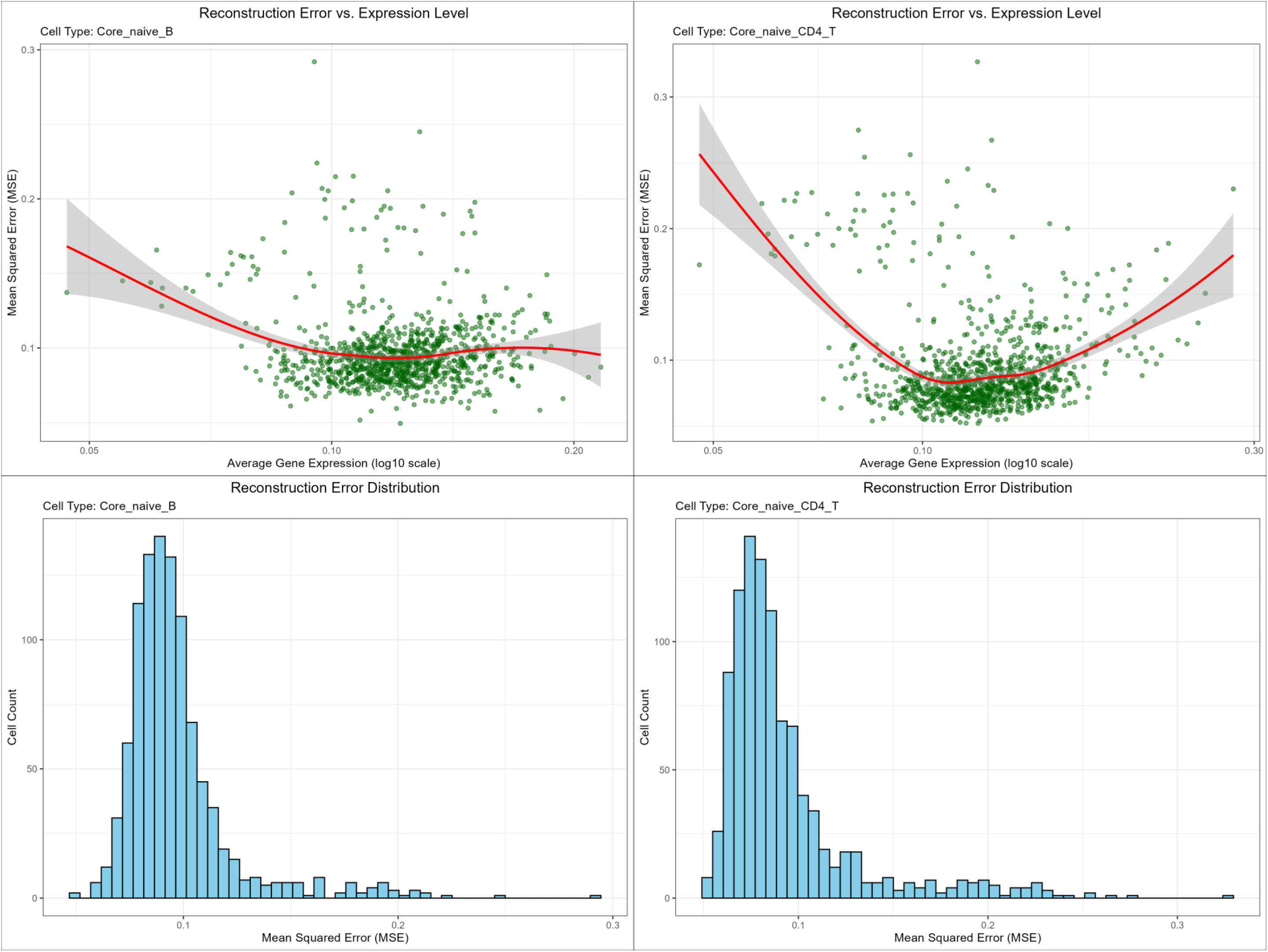
Reconstruction error analysis of the Hierarchical Deep Neural Network model. (Top row) Scatter plots showing the relationship between reconstruction error (mean squared error, MSE) and average gene expression level (log10 scale) for core naive B cells (left) and core naive CD4+ T cells (right). (Bottom row) Histograms showing the distribution of reconstruction error (mean squared error, MSE) across all analyzed genes for core naive B cells (left) and core naive CD4+ T cells (right).

**Figure 13.**
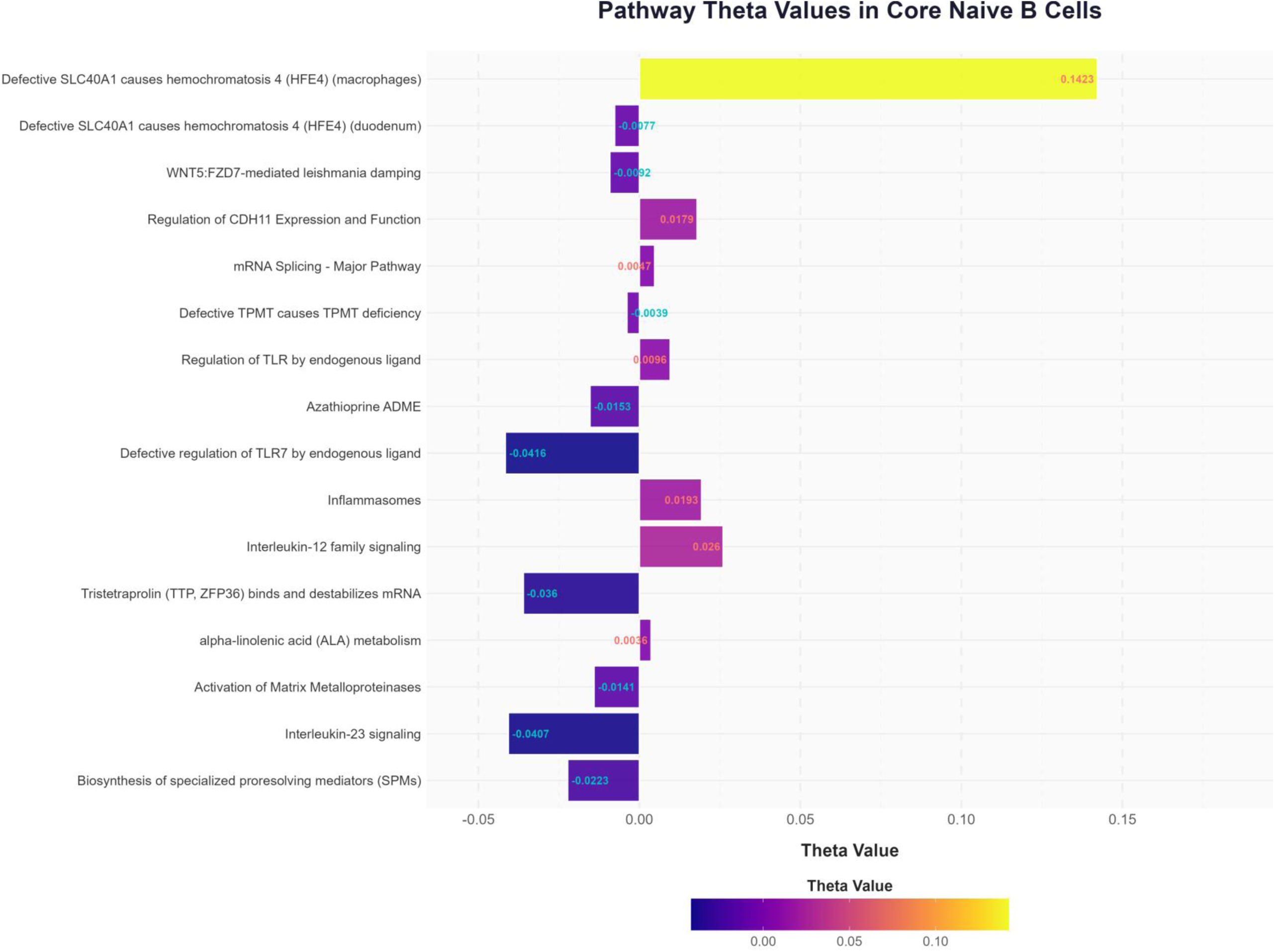
Cell type-specific causal effect quantification of 16 RA-related signaling pathways from the Reactome database. The heatmap shows the causal effect size (Theta value) of each pathway across immune cell types, estimated by the validated single-cell DML model. Color intensity represents the magnitude of the causal effect, with red indicating a positive causal effect on RA and blue indicating a negative causal effect.

**Figure 14.**
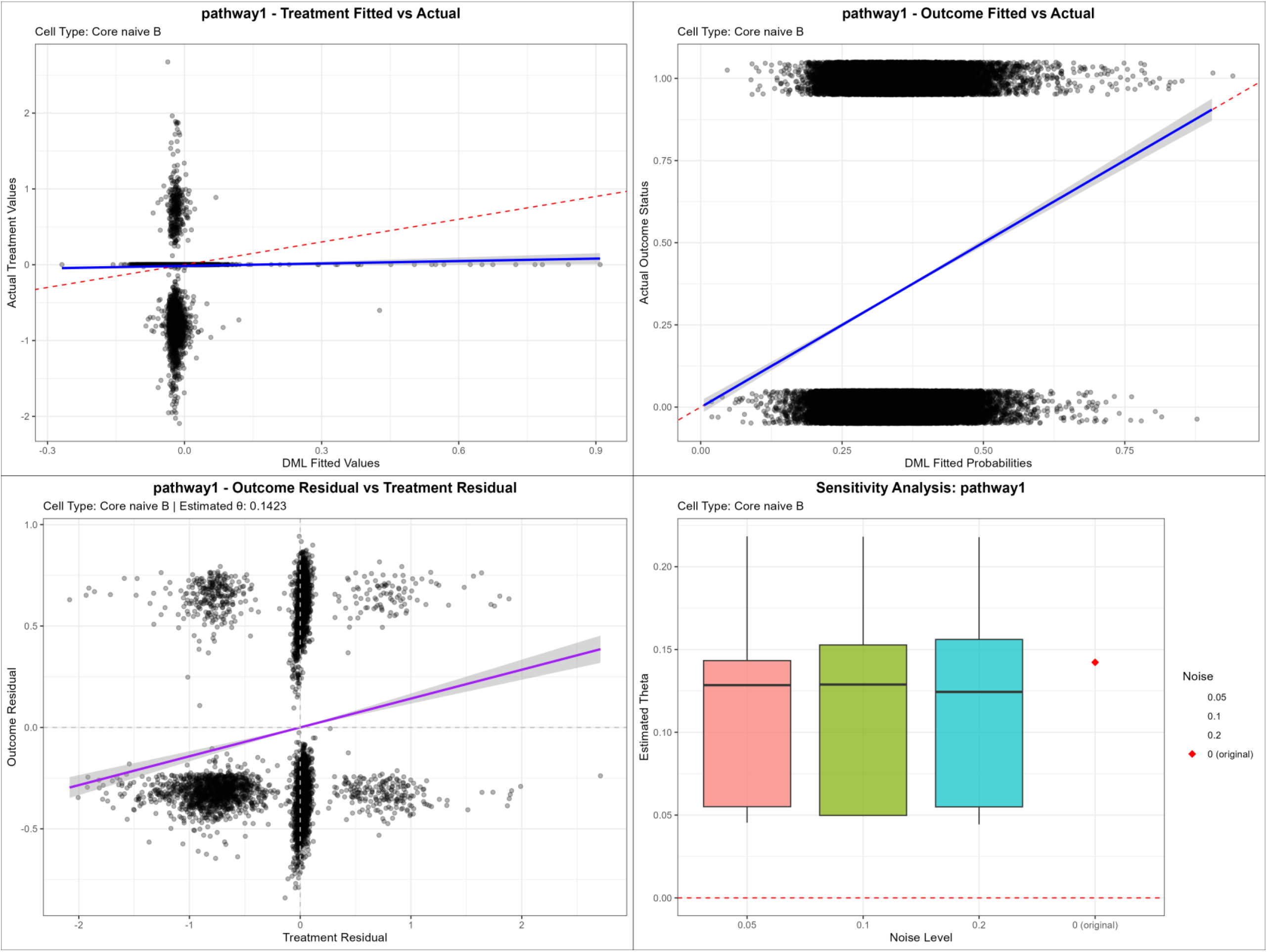
Residual diagnostic plots and sensitivity analysis for pathway1 in Core naive B cells (Top left) Scatter plot of actual treatment values against DML fitted values for pathway1, with the blue solid line representing the fitted regression line and the red dashed line indicating the reference line of perfect fit. (Top right) Scatter plot of actual outcome status against DML fitted probabilities for pathway1, with the blue solid line representing the fitted regression line and the red dashed line indicating the reference line of perfect fit. (Bottom left) Scatter plot illustrating the association between outcome residuals and treatment residuals for pathway1, with the purple solid line indicating the linear regression fit, the grey shaded area denoting the 95% confidence interval, and the estimated treatment effect θ = 0.1423. (Bottom right) Sensitivity analysis of the estimated treatment effect θ for pathway1 under varying noise levels: boxplots display the distribution of θ estimates at 0.05, 0.1, and 0.2 noise levels, the red diamond marks the original estimate at 0 noise level, and the red dashed line indicates the null effect threshold (θ = 0).

After Bonferroni correction, the correlation in Core naive B cells yielded an adjusted p-value of 5.136×10⁻⁴ (0.00051, calculated as 3.21×10⁻⁵ × 16), which was statistically significant (p < 0.05).

## Discussion

Our research demonstrates that statistical biology and systems biology can converge on the same biological truth. The results from large-scale population studies (GWAS + eQTL + Transcriptome-wide Mendelian Randomization - TWMR) show a significant connection with the findings from single-cell transcriptomics of human samples analyzed via deep learning coupled with Double Machine Learning (DML) for causal inference. This cross-scale unity suggests that the biological principles governing macro (population) and micro (single-cell) levels exhibit consistency. This likely stems from the conservation of certain core mechanistic elements across scales.

## Key Implications

1. Transcriptome-wide Mendelian Randomization (TWMR) based on human population data serves as the gold standard for validation. Single-cell transcriptomics is directly obtained from human samples (such as surgical specimens and tissues of patients), and the two are significantly correlated, which can substantially narrow the Translational Gap.:This brings research findings closer to actual human physiology and pathology. A major reason for the failure of biomedical research translation (“bench-to-bedside”) is the discrepancy between human tissue biology and experimental models (cell lines, animal models). TWMR, based on human population data, completely avoids issues like species bias or lack of genetic diversity inherent in animal/cell models, reflecting true human genetic-disease associations. Single-cell transcriptomics provides direct molecular observations from human samples (e.g., patient surgical specimens, tissues), avoiding the problem of model systems failing to replicate human cellular heterogeneity. Their agreement indicates high consistency between population-level statistical discoveries (free of model bias) and molecular-level observations in human cells (free of species bias), dramatically shortening the translational pathway from basic research to human disease.
2. Unsupervised deep learning-based data compression combined with orthogonalized double machine learning for causal inference may be an effective approach to achieve data alignment between statistical biology and systems biology across different scales: Molecular drivers identified at the single-cell level demonstrably correlate with phenotypes at the population level. Population statistical trends can be directly mapped to cellular molecular mechanisms, reducing the absolute need for intermediate model system validation (animal or cell models). This cross-scale integrated validation provides strong support for building a unified theoretical framework connecting molecules to phenotypes. This integration will drastically shorten the path from observational studies to mechanistic verification and accelerate translation from basic research to clinical applications. It effectively mitigates the challenges reductionism faces when addressing complex diseases. This discovery will drive a shift in biomedical research from reliance on “animal/cell models” towards being “directly driven by human samples.” Furthermore, this cross-scale consistency suggests that rare disease research can directly utilize single-cell data from patient cases, alleviating the critical limitation of insufficient sample sizes for population GWAS in rare diseases.

At present, remarkable progress has been made in the field combining single-cell RNA sequencing (scRNA-seq) with gene perturbation tools (such as in silico gene knockout). Single-cell technologies enable the investigation of signal transduction in complex tissues at unprecedented resolution. Gene knockout (KO) experiments are a well-validated and powerful approach for gene function research. However, systematic KO experiments targeting a large number of genes are often difficult to implement due to limited experimental and animal resources. Several important biological macromolecules and molecular networks have been identified using in silico gene perturbation. Using a deep generative model, Sheban F et al. constructed a gene perturbation network, mapped a detailed roadmap of the gene circuit in tumor-associated macrophages (TAMs), and identified ZEB2 as a master switch with therapeutic potential^[4]^. The pathogenesis of Parkinson’s disease (PD) is closely related to neuroinflammation, and type I interferon (IFN-I) plays a key role in regulating immune and inflammatory responses. However, the cell type-specific characteristics of IFN and its underlying mechanisms in PD have not been fully characterized. In silico gene perturbation confirmed that NFATc2 regulates the IFN-I response and neuroinflammation. In subsequent experiments using a 1-methyl-4-phenylpyridinium (MPP+)-induced BV2 cell model, inhibition of NFATc2 resulted in decreased IFN-β levels, impaired signal transducer and activator of transcription 1 (STAT1) phosphorylation, and attenuated activation of the nuclear factor kappa-B (NF-κB) pathway. Furthermore, downregulation of NFATc2 alleviated the adverse effects on SH-SY5Y cells co-cultured in conditioned medium^[5]^. Capillary malformation (CM) is a congenital vascular anomaly affecting the skin, mucosa, and brain, yet the understanding of its vascular pathogenesis remains limited. Nguyen V et al. applied spatial whole-transcriptome analysis (GeoMx) and gene set enrichment analysis (GSEA) to CM lesions at the single-vessel level. Differentially expressed genes were validated by immunofluorescence staining. Phosphatasomics analysis was performed to reveal lesion-wide phosphorylation sites on proteins. scRNA-seq was conducted on CM-derived induced pluripotent stem cells (iPSCs) to determine the differentiation trajectory of lesional vascular lineages. In silico gene perturbation was used to predict candidate genes regulating the progression of vascular pathology, followed by functional validation using a Tet-on system in CM iPSC-derived endothelial cells (ECs) ^[6]^.

These advances are mainly attributed to the development of efficient computational tools, including scTenifoldKnk, GenKI, scLAMBDA, and GPerturb.

scLAMBDA is a deep generative learning framework designed to model and predict the transcriptional responses of single cells to genetic perturbations, including single-gene and combinatorial multi-gene perturbations. By leveraging gene embeddings from large language models, scLAMBDA effectively integrates prior biological knowledge and decouples the basal cell state from perturbation-specific signature representations. Through comprehensive evaluation on multiple single-cell CRISPR perturbation sequencing datasets, scLAMBDA consistently outperforms state-of-the-art methods in predicting perturbation outcomes, achieving higher prediction accuracy^[7]^.

ScTenifoldKnk is an efficient in silico KO tool that enables systematic gene function KO studies using scRNA-seq data.In scTenifoldKnk analysis, a gene regulatory network (GRN) is first constructed from scRNA-seq data of wild-type (WT) samples, followed by virtual deletion of the target gene from the constructed GRN. Manifold alignment is applied to align the perturbed GRN with the original GRN to identify differentially regulated genes, which are used to infer the function of the target gene in the analyzed cells. Osorio D et al. demonstrated that scTenifoldKnk-based in silico KO analysis recapitulated the major findings of real animal KO experiments and recovered the expected functions of genes in relevant cell types^[8]^.

Hu Y et al. developed CytoTalk, a tool for de novo construction of cell type-specific signaling networks using single-cell transcriptomic data. CytoTalk takes integrated intracellular and extracellular gene networks as input and identifies candidate pathways using the award-winning Steiner Forest algorithm. By combining high-throughput spatial transcriptomic data and scRNA-seq data with receptor gene perturbation, the researchers demonstrated that CytoTalk offers significant improvements over existing algorithms. To better understand the plasticity of signaling networks across tissues and developmental stages, a comparative analysis of signaling networks between macrophages and endothelial cells in human adult and fetal tissues was performed. The analysis revealed enhanced overall plasticity of signaling networks in adult tissues, as well as specific network nodes that contribute to this enhanced plasticity. CytoTalk enables de novo construction of signal transduction pathways and facilitates comparative analysis of these pathways across tissues and disease states^[9]^.

Gene Knockout Inference (GenKI) is an in silico KO tool for gene function prediction using scRNA-seq data when only wild-type (WT) samples are available. GenKI is designed to capture the patterns of gene regulatory changes induced by KO perturbation in an unsupervised manner, and provides a robust and scalable framework for gene function research via its variational graph autoencoder (VGAE) model. To achieve this, GenKI adopts a VGAE model to learn the latent representations of genes and gene-gene interactions from the input WT scRNA-seq data and the derived single-cell gene regulatory network (scGRN). In silico KO data are generated by computationally removing all edges of the target gene to be knocked out for functional studies from the scGRN. Differences between WT and in silico KO data are distinguished using the corresponding latent parameters derived from the trained VGAE model. Simulations showed that GenKI accurately approximates the perturbation profile upon gene KO and outperforms state-of-the-art techniques under a range of evaluation conditions. Using publicly available scRNA-seq datasets, the researchers demonstrated that GenKI recapitulates findings from real animal KO experiments and accurately predicts cell type-specific functions of KO genes. Therefore, GenKI provides an in silico alternative to KO experiments, and may partially replace the reliance on genetically modified animals or other gene perturbation systems^[10]^.

GPerturb is a Gaussian process sparse perturbation regression model specifically designed for single-cell CRISPR screening/perturbation data. One of its core functions is the accurate prediction of gene expression levels after perturbation, which has been validated using 6 real-world publicly available standard datasets of single-cell perturbation^[11]^.

The validity and accuracy of these in silico gene knockout or perturbation models are mainly verified through the following approaches: 1. Direct comparison of model predictions with published KO results from real animal or cell experiments. 2. Functional validation of model-predicted outputs, such as key genes or regulators, via subsequent independent animal or cell experiments. 3. Consistency of model predictions with well-established biological knowledge, including conformity to known biological mechanisms or pathways. In summary, the validity and accuracy of these models are mainly verified by comparison with real experimental data, validation via subsequent independent experiments, and consistency with known biological knowledge. These gene perturbation tools reduce the demand for animal experiments. However, there are currently no reports of direct validation of these computational tools for in silico gene perturbation using results from large-scale population-based genome-wide association study (GWAS), expression quantitative trait locus (eQTL), and Transcriptome-wide Mendelian Randomization (TWMR) analyses. The advantage of Double Machine Learning (DML) for estimating causal effects in single-cell transcriptomics lies in its Neyman orthogonality, which may render DML less susceptible to model specification. DML is first-order insensitive to the error of the model itself.

Analysis of 16 signaling pathways from the Reactome database was performed using the validated model. The results indicated that the pathway Defective SLC40A1 causes hemochromatosis 4 (HFE4) (macrophages) had the highest effect size and contained only two genes, ceruloplasmin (CP) and SLC40A1. We reviewed relevant literature to validate the findings from the model calculation.

Pilling LC et al. compared the prevalence, incidence, and mortality between carriers of the HFE p.C282Y genetic variant (which causes the majority of hereditary hemochromatosis type 1 cases) and non-carriers in a large community-based sample of European ancestry from the UK Biobank cohort study (451,243 European ancestry volunteers). A significantly increased risk of rheumatoid arthritis (RA) was found in carriers (odds ratio [OR] 2.23, 95% confidence interval [CI] 1.51 to 3.31, P<0.001) ^[12]^. Barton JC et al. conducted a retrospective study of autoimmune conditions (ACs) at the time of diagnosis in 235 probands with hemochromatosis. The prevalence of Hashimoto’s thyroiditis, rheumatoid arthritis, and ankylosing spondylitis was 8.1% (95% CI: 5.1 to 12.5), 1.7% (95% CI: 0.6 to 4.6), and 0.0085% (95% CI: 0.0015 to 0.0337), respectively. They concluded that ACs are common in hemochromatosis probands, especially women and probands with a history of ACs in first-degree family members^[13]^. Carini M et al. analyzed the frequency of the p.Cys282Tyr mutation and the -174G>C polymorphism of the IL-6 gene in patients affected by autoimmune diseases, including rheumatoid arthritis (RA) and systemic lupus erythematosus (SLE), and found that the overall frequency of the p.Cys282Tyr mutation was significantly reduced in patients with RA and SLE^[14]^. Nauffal V et al. developed a machine learning model to measure native myocardial T1 time (a marker of myocardial fibrosis) in 41,505 UK Biobank participants who underwent cardiac magnetic resonance imaging. Prolonged T1 time was associated with diabetes mellitus, nephropathy, aortic stenosis, cardiomyopathy, heart failure, atrial fibrillation, conduction disorders, and rheumatoid arthritis. Genome-wide association analysis identified 11 independent loci associated with T1 time. The identified loci involved genes related to glucose transport (SLC2A12), iron homeostasis (HFE, TMPRSS6), tissue repair (ADAMTSL1, VEGFC), oxidative stress (SOD2), cardiac hypertrophy (MYH7B), and calcium signaling (CAMK2D) ^[15]^. In addition, we identified multiple case reports of rheumatoid arthritis associated with hemochromatosis, none of which performed genotyping for the homozygous C282Y mutation^[16]^ ^[17]^ ^[18][19] [20] [21] [22] [23] [24] [25]^.

The Reactome pathway Defective SLC40A1 causes hemochromatosis 4 (HFE4) (macrophages) comprises only two genes, SLC40A1 and CP. SLC40A1 is listed as a marker gene for ferroptosis and is involved in iron metabolism and oxidative stress in RA^[26]^. Loss-of-function (LoF) mutations in SLC40A1 lead to defective iron export function of ferroportin 1 (FPN1) in macrophages, which is the sole core pathogenic mechanism of macrophage-involved HFE4. In the inflammatory milieu of rheumatoid arthritis (RA), macrophages that still display an “anti-inflammatory M2-type” phenotype based on surface markers (such as the M2 markers CD14 and CD163) have actually lost their anti-inflammatory activity and switched to a pro-inflammatory phenotype, accompanied by downregulated SLC40A1 expression. The classic M1/M2 phenotypic classification cannot accurately reflect the actual function of macrophages in inflammatory diseases^[27]^. In studies of juvenile rheumatoid arthritis (JRA), plasma ceruloplasmin concentrations were found to be significantly reduced^[28]^. In studies of sporadic rheumatoid arthritis and animal experimental studies, ceruloplasmin has been described as a marker of oxidative stress and inflammation, with significant alterations in RA^[29]^ ^[30]^ ^[31]^ ^[32]^ ^[33]^ ^[34] [35] [36] [37] [38] [39] [40] [41] [42] [43] [44] [45] [46] [47] [48] [49].^

Finally, in pharmacological studies, auranofin (AUR), an anti-rheumatoid arthritis drug, has been shown to potently upregulate the expression of hepatic hepcidin. The researchers unexpectedly found that canonical iron regulatory signaling pathways, including the BMP/SMAD and IL-6/JAK2/STAT3 pathways, play an indispensable role in mediating the effects of AUR. The researchers suggested that AUR is a novel activator of hepatic hepcidin expression and iron depletion via distinct mechanisms, and may have potential for the treatment of hemochromatosis and diseases related to hepcidin deficiency^[50]^.

Hereditary hemochromatosis type 1 is a monogenic disorder. The above studies on hemochromatosis may suggest that there is a genuine and strong association between the CP gene and rheumatoid arthritis.

## Our study has limitations

1. The population GWAS cohort comprised 460,000 individuals, which may still yield an insufficient number of rheumatoid arthritis (RA) cases. TWMR has limited sensitivity to rare variants.
2. The single-cell dataset included only 11 RA samples. Data sparsity and batch effects in scRNA-seq could impact model accuracy.
3. Computational constraints prevented running analyses on all genes or performing extensive independent replications.
4. To date, we have verified a Pearson correlation between the results of Transcriptome-wide Mendelian Randomization (TWMR) and those of causal inference from deep learning combined with double machine learning on single-cell transcriptomes in only one complex disease, namely rheumatoid arthritis. It remains to be further investigated whether this correlation is present in other complex diseases.

## Future Outlook

Our work explores the possibility that GWAS (population-scale) and single-cell studies (cellular-scale), though operating at different levels, exhibit significant overlap or consistency. The basis for this overlap may lie in the conservation of core biological mechanisms across scales. However, the extent and boundaries of this overlap, particularly with larger-scale GWAS and single-cell datasets, remain undefined and require further investigation. Moreover, the fundamental reason *why* these core mechanisms are conserved across scales currently lacks a clear explanation using reductionist methods.

Should future research confirm that this cross-scale consistency is very high, it would be immensely beneficial for establishing and validating systems biology models built directly on human single-cell data. Despite technological advances like single-cell omics, understanding complex human diseases remains challenging (e.g., Alzheimer’s disease (AD) “remains elusive”). A key debate centers on why systems biology approaches (e.g., integrating gene regulatory networks) are “largely untapped” in AD research, leading to a focus on single proteins or mechanisms, ignoring the disease as a complex system^[51][52]^.

Utilizing systems biology models built from large-scale human single-cell data, validated and calibrated with large-scale GWAS results, offers the potential to establish reference models. Supported by supercomputing capabilities, such standardized human systems biology reference models could computationally quantify the effect sizes of genes and pathways. This would allow establishing a standardized, quantitative frame of reference (“reference system”) for each disease or trait. Findings from diverse animal and cell experimental models could then be mapped onto this common reference system. This approach addresses the current lack of holistic perspective (“blind men and the elephant” problem) in biomedical research. Ultimately, it promises to dramatically accelerate our understanding of complex diseases and enhance the efficiency of clinical translation.

## Data Availability

All data generated during this study are available from the authors upon reasonable request.

## References

[1] Fischer DS, Villanueva MA, Winter PS, Shalek AK. Adapting systems biology to address the complexity of human disease in the single-cell era. Nat Rev Genet. 2025;26(8):514–531. doi:10.1038/s41576-025-00821-6

[2] Jiang L, Zheng Z, Fang H, Yang J. A generalized linear mixed model association tool for biobank-scale data. Nat Genet. 2021;53(11):1616–1621. doi:10.1038/s41588-021-00954-4

[3] He Z, Glass MC, Venkatesan P, et al. Progression to rheumatoid arthritis in at-risk individuals is defined by systemic inflammation and by T and B cell dysregulation. Sci Transl Med. 2025;17(817):eadt7214. doi:10.1126/scitranslmed.adt7214

[4] Sheban F, Phan TS, Xie K, et al. ZEB2 is a master switch controlling the tumor-associated macrophage program. Cancer Cell. 2025;43(7):1227–1241.e11. doi:10.1016/j.ccell.2025.03.021

[5] Quan P, Li X, Si Y, et al. Single cell analysis reveals the roles and regulatory mechanisms of type-I interferons in Parkinson’s disease. Cell Commun Signal. 2024;22(1):212. Published 2024 Apr 2. doi:10.1186/s12964-024-01590-1

6. Nguyen V, Mao I, He S, et al. Single-vessel transcriptome map pathological landscapes and reveal NR2F2-mediated smooth muscle cell phenotype acquisition in capillary malformations. Preprint. bioRxiv. 2025;2025.09.02.673874. Published 2025 Sep 17. doi:10.1101/2025.09.02.673874

[7] Wang G, Liu T, Zhao J, Cheng Y, Zhao H. Modeling and predicting single-cell multi-gene perturbation responses with scLAMBDA. Preprint. bioRxiv. 2024;2024.12.04.626878. Published 2024 Dec 8. doi:10.1101/2024.12.04.626878

[8] Osorio D, Zhong Y, Li G, et al. scTenifoldKnk: An efficient virtual knockout tool for gene function predictions via single-cell gene regulatory network perturbation. Patterns (N Y). 2022;3(3):100434. Published 2022 Feb 1. doi:10.1016/j.patter.2022.100434

[9] Hu Y, Peng T, Gao L, Tan K. CytoTalk: De novo construction of signal transduction networks using single-cell transcriptomic data. Sci Adv. 2021;7(16):eabf1356. Published 2021 Apr 14. doi:10.1126/sciadv.abf1356

[10] Yang Y, Li G, Zhong Y, et al. Gene knockout inference with variational graph autoencoder learning single-cell gene regulatory networks. Nucleic Acids Res. 2023;51(13):6578–6592. doi:10.1093/nar/gkad450

[11] Xing H, Yau C. GPerturb: Gaussian process modelling of single-cell perturbation data. Nat Commun. 2025;16(1):5423. Published 2025 Jul 1. doi:10.1038/s41467-025-61165-7

[12] Pilling LC, Tamosauskaite J, Jones G, et al. Common conditions associated with hereditary haemochromatosis genetic variants: cohort study in UK Biobank. BMJ. 2019;364:k5222. Published 2019 Jan 16. doi:10.1136/bmj.k5222

[13] Barton JC, Barton JC. Autoimmune Conditions in 235 Hemochromatosis Probands with HFE C282Y Homozygosity and Their First-Degree Relatives. J Immunol Res. 2015;2015:453046. doi:10.1155/2015/453046

[14] Carini M, Fredi M, Cavazzana I, et al. Frequency Evaluation of the Interleukin-6 -174G>C Polymorphism and Homeostatic Iron Regulator (HFE) Mutations as Disease Modifiers in Patients Affected by Systemic Lupus Erythematosus and Rheumatoid Arthritis. Int J Mol Sci. 2023;24(22):16300. Published 2023 Nov 14. doi:10.3390/ijms242216300

[15] Nauffal V, Di Achille P, Klarqvist MDR, et al. Genetics of myocardial interstitial fibrosis in the human heart and association with disease. Nat Genet. 2023;55(5):777–786. doi:10.1038/s41588-023-01371-5

[16] Harada M, Honma Y, Shiba E, Tomosugi N, Harada R. Tocilizumab, a Humanized Anti-interleukin-6 Receptor Antibody, Induces Hepatic Iron Overload in a Susceptible Patient. Intern Med. 2025;64(9):1334–1337. doi:10.2169/internalmedicine.4329-24

[17] Chit Su HM, Putchakayala K. A Mystery of Joint Pain: Is It Rheumatoid Arthritis (RA) or Hereditary Hemochromatosis (HH)?. Cureus. 2022;14(12):e33037. Published 2022 Dec 28. doi:10.7759/cureus.33037

[18] Barbosa FB, Callegari A, Sarinho JC, Lucena J, Casagrande R, de Souza BD. Hemochromatosis simulating rheumatoid arthritis: a case report. Rev Bras Reumatol. 2014;54(1):62–64.

19. Zafar S, Badsha H. Atypical arthritis due to combined hereditary hemochromatosis and active hepatitis C. J Clin Rheumatol. 2008;14(1):21–23. doi:10.1097/RHU.0b013e31816373f3

[20] Wernicke D, Seipelt E, Schmidt WA, Gromnica-Ihle E. Manifestation of rheumatoid arthritis in a patient with hereditary haemochromatosis. Rheumatol Int. 2006;26(10):939–941. doi:10.1007/s00296-006-0113-8

[21] Schumacher HR Jr. Arthropathy in hemochromatosis. Hosp Pract (1995). 1998;33(3):81–94. doi:10.1080/21548331.1998.11443654

[22] Bensen WG, Laskin CA, Little HA, Fam AG. Hemochromatotic arthropathy mimicking rheumatoid arthritis. A case with subcutaneous nodules, tenosynovitis, and bursitis. Arthritis Rheum. 1978;21(7):844–848. doi:10.1002/art.1780210717

[23] Ito K, Minamimoto R, Morooka M, Kubota K. A case of secondary hemochromatosis with high uptake of liver in F-18 FDG PET/CT imaging. Clin Nucl Med. 2011;36(7):606–608. doi:10.1097/RLU.0b013e318217ae7c

[24] Lonardo A, Neri P, Mascia MT, Pietrangelo A. Hereditary hemochromatosis masquerading as rheumatoid arthritis. Ann Ital Med Int. 2001;16(1):46–49.

[25] Espinosa-Morales R, Escalante A. Diagnostic confusion caused by hepatitis C: hemochromatosis presenting as rheumatoid arthritis. J Rheumatol. 1998;25(12):2459–2463.

[26] Xia P, Du H, Ren Y, Zhang Q, Shi Y. Machine learning reveals ferroptosis and therapeutic targets in rheumatoid arthritis. Medicine (Baltimore). 2025;104(39):e44384. doi:10.1097/MD.0000000000044384

[27] Quero L, Hanser E, Manigold T, Tiaden AN, Kyburz D. TLR2 stimulation impairs anti-inflammatory activity of M2-like macrophages, generating a chimeric M1/M2 phenotype. Arthritis Res Ther. 2017;19(1):245. Published 2017 Nov 2. doi:10.1186/s13075-017-1447-1

[28] Ashour M, Salem S, Hassaneen H, et al. Antioxidant status in children with juvenile rheumatoid arthritis (JRA) living in Cairo, Egypt. Int J Food Sci Nutr. 2000;51(2):85–90. doi:10.1080/096374800100787

[29] Strecker D, Mierzecki A, Radomska K. Copper levels in patients with rheumatoid arthritis. Ann Agric Environ Med. 2013;20(2):312–316.

[30] Honkanen V, Konttinen YT, Sorsa T, et al. Serum zinc, copper and selenium in rheumatoid arthritis. J Trace Elem Electrolytes Health Dis. 1991;5(4):261–263.

[31] Stojan B, Hasler F. Immunologische Befunde im Serum und in der Gelenkflüssigkeit bei Patienten mit rheumatoider Arthritis [Immunological findings in serum and synovial fluid in patients with rheumatoid arthritis (author’s transl)]. Wien Klin Wochenschr. 1977;89(21):713–716.

[32] Ben-Hadj-Mohamed M, Khelil S, Ben Dbibis M, et al. Hepatic Proteins and Inflammatory Markers in Rheumatoid Arthritis Patients. Iran J Public Health. 2017;46(8):1071–1078.

[33] Ahmadzadeh N, Shingu M, Nobunaga M, Yasuda M. Correlation of metal-binding proteins and proteinase inhibitors with immunological parameters in rheumatoid synovial fluids. Clin Exp Rheumatol. 1990;8(6):547–551.

[34] Li TW, Zheng BR, Huang ZX, et al. Screening disease-associated proteins from sera of patients with rheumatoid arthritis: a comparative proteomic study. Chin Med J (Engl). 2010;123(5):537–543.

[35] Sokolov AV, Acquasaliente L, Kostevich VA, et al. Thrombin inhibits the anti-myeloperoxidase and ferroxidase functions of ceruloplasmin: relevance in rheumatoid arthritis. Free Radic Biol Med. 2015;86:279–294. doi:10.1016/j.freeradbiomed.2015.05.016

[36] Guan X, Huang Z, Chen J, Fan X, Zheng SG. The double-edged sword role of copper in rheumatoid arthritis: Mechanisms, therapeutics, and challenges. J Autoimmun. 2025;157:103484. doi:10.1016/j.jaut.2025.103484

[37] Matsumoto Y, Sugioka Y, Tada M, et al. Impact of disease burden or inflammation on nutritional assessment by the GLIM criteria in female patients with rheumatoid arthritis. Clin Nutr ESPEN. 2022;52:353–359. doi:10.1016/j.clnesp.2022.09.016

[38] Shan L, Tong L, Hang L, Fan H. Fangchinoline supplementation attenuates inflammatory markers in experimental rheumatoid arthritis-induced rats. Biomed Pharmacother. 2019;111:142–150. doi:10.1016/j.biopha.2018.12.043

[39] Dai W, Qi C, Wang S. Synergistic effect of glucosamine and vitamin E against experimental rheumatoid arthritis in neonatal rats. Biomed Pharmacother. 2018;105:835–840. doi:10.1016/j.biopha.2018.05.136

[40] Wang S, Tian S, Li M, Li Z. Methionine attenuates the intensity of rheumatoid arthritis by downregulating NF-κB and iNOS expression in neonatal rats. 3 Biotech. 2018;8(7):303. doi:10.1007/s13205-018-1311-2

[41] Sahebari M, Ayati R, Mirzaei H, et al. Serum Trace Element Concentrations in Rheumatoid Arthritis. Biol Trace Elem Res. 2016;171(2):237–245. doi:10.1007/s12011-015-0501-6

[42] Sultana F, Rasool M. A novel therapeutic approach targeting rheumatoid arthritis by combined administration of morin, a dietary flavanol and non-steroidal anti-inflammatory drug indomethacin with reference to pro-inflammatory cytokines, inflammatory enzymes, RANKL and transcription factors. Chem Biol Interact. 2015;230:58–70. doi:10.1016/j.cbi.2015.02.007

[43] Meki AR, Hamed EA, Ezam KA. Effect of green tea extract and vitamin C on oxidant or antioxidant status of rheumatoid arthritis rat model. Indian J Clin Biochem. 2009;24(3):280–287. doi:10.1007/s12291-009-0053-7

[44] Alorainy M. Effect of allopurinol and vitamin e on rat model of rheumatoid arthritis. Int J Health Sci (Qassim). 2008;2(1):59–67.

[45] Isik A, Koca SS, Ustundag B, Celik H, Yildirim A. Paraoxonase and arylesterase levels in rheumatoid arthritis. Clin Rheumatol. 2007;26(3):342–348. doi:10.1007/s10067-006-0300-8

[46] Nagler RM, Salameh F, Reznick AZ, Livshits V, Nahir AM. Salivary gland involvement in rheumatoid arthritis and its relationship to induced oxidative stress. Rheumatology (Oxford). 2003;42(10):1234–1241. doi:10.1093/rheumatology/keg362

[47] Cogalgil S, Taysi S. Levels of antioxidant proteins and soluble intercellular adhesion molecule-1 in serum of patients with rheumatoid arthritis. Ann Clin Lab Sci. 2002;32(3):264–270.

[48] Taysi S, Polat F, Gul M, Sari RA, Bakan E. Lipid peroxidation, some extracellular antioxidants, and antioxidant enzymes in serum of patients with rheumatoid arthritis. Rheumatol Int. 2002;21(5):200–204. doi:10.1007/s00296-001-0163-x

[49] Louro MO, Cocho JA, Mera A, Tutor JC. Immunochemical and enzymatic study of ceruloplasmin in rheumatoid arthritis. J Trace Elem Med Biol. 2000;14(3):174–178. doi:10.1016/S0946-672X(00)80007-3

[50] Yang L, Wang H, Yang X, et al. Auranofin mitigates systemic iron overload and induces ferroptosis via distinct mechanisms. Signal Transduct Target Ther. 2020;5(1):138. Published 2020 Jul 31. doi:10.1038/s41392-020-00253-0

[51] Rahimzadeh N, Srinivasan SS, Zhang J, Swarup V. Gene networks and systems biology in Alzheimer’s disease: Insights from multi-omics approaches. Alzheimers Dement. 2024;20(5):3587–3605. doi:10.1002/alz.13790

[52] Rollo JL, Banihashemi N, Vafaee F, Crawford JW, Kuncic Z, Holsinger RM. Unraveling the mechanistic complexity of Alzheimer’s disease through systems biology. Alzheimers Dement. 2016;12(6):708–718. doi:10.1016/j.jalz.2015.10.010

